# Deceased donor kidney degradomics indicates cytoskeletal proteolytic alterations impacting post-transplant function

**DOI:** 10.1101/2021.06.10.21254305

**Authors:** Rebecca H. Vaughan, Jean-Claude Kresse, Louise K. Farmer, Marie L. Thézénas, Benedikt M. Kessler, Jan H. N. Lindeman, Edward J Sharples, Gavin I. Welsh, Rikke Nørregaard, Rutger J. Ploeg, Maria E. Kaisar

## Abstract

**Background:** In brain death, cerebral injury contributes to systemic biological dysregulation, causing significant cellular stress in donor kidneys that adversely impacts the quality of grafts. Here, we hypothesized that DBD kidneys may undergo proteolytic processes that renders grafts susceptible to post-transplant dysfunction.

**Material & Methods:** Using mass spectrometry and immunoblotting analyses, we profiled degradation patterns of cytoskeletal proteins in deceased (n=55) and living (n=10) donor kidneys.

**Results:** We found that in DBD kidneys, key podocyte cytoskeletal proteins had been proteolytically cleaved. Generated degradation profiles were not associated to donor related demographic and clinical factors but were associated to suboptimal post-transplant function. Strikingly, α-actinin -4 and Talin-1 degradation profiles were not observed in circulatory-death or living-donor kidneys. As Talin-1 is a specific proteolytic target of Calpain-1, we investigated a potential trigger of Calpain activation and Talin-1 degradation using ex-vivo precision-cut human kidney slices and in-vitro immortalised human podocytes. Notably, we found that Transforming-Growth Factor-β (TGF-β) activated Calpain-1 and proteolytically cleaved Talin-1 to generate distinct peptide fragments. These peptide fragments were of similar size and matched the degradation patterns observed in DBD kidneys. Talin-1 degradation was prevented in vitro by Calpain-1 inhibition.

**Conclusions:** Here, we provide initial evidence that DBD kidneys are susceptible to cytoskeletal protein degradation that impacts posttransplant kidney function. Subsequent studies should aim to further investigate the link between brain death and activation of proteolytic pathways exploring new therapeutic opportunities.

## 1. INTRODUCTION

Kidney transplantation is the only curative treatment for patients with end-stage kidney disease. When compared to dialysis, transplantation profoundly increases life-expectancy, improves quality of life (QoL) and is cost-effective ^1–3^.

Deceased kidney donation after brain death (DBD) is the main source of transplants, yet these grafts yield inferior short and long-term transplant outcomes when compared to living donation. This makes kidney allograft failure one of the most common morbidities to start dialysis in the UK and the USA ^4^.

The process of brain death and subsequent donor management in intensive care units is associated with a complex homeostatic dysregulation in the donor ^5, 6^ strongly suggesting that brain death negatively impacts organ quality ^7, 8^. In fact, brain death associates with a specific pattern of damage to renal substructures including the podocyte, which will compromise post-transplant graft function and graft survival ^9–15^. As yet, the molecular processes underlying brain death-associated glomerular injury, with the potential for targeted interventions and pharmaceutical prevention, are unknown^16^.

Proteolytic damage of podocytes is a common effector pathway for glomerular diseases^17–19^. Recent studies in experimental models, suggest that the maintenance of renal filtration barrier largely relies on proteases as key regulators of podocyte integrity and kidney function ^20^. Evidence that dysregulation of proteolytic pathways is a key driver for human disease signifying proteases as promising targets for tailored therapeutics^21^.

In this study, we hypothesized that brain death induced kidney cytoskeletal alterations mediated by proteolytic processes are associated with subsequent suboptimal transplant outcomes. To investigate this, we analysed donor kidney biopsies that were selected on the basis of paired transplant outcomes and classified the donors as suboptimal (SO) or good (GO) subgroups. We initially profiled the protein degradome (proteolytic modifications) of pre-implantation kidney biopsies from DBD grafts with contrasting long-term functional outcomes by utilizing the Protein Topography and Migration Analysis Platform (PROTOMAP)^22^ with a focus on proteins enriched in the kidney cytoskeleton. Next, to investigate whether protein degradation was donor-type specific, we profiled the proteolytic patterns of a number of key proteins on a separate cohort of DBD, DCD and living donor kidney biopsies by immunoblotting that allowed detailed description of proteolytic fragments. Based on our previous research findings,^23, 24^ and to gather initial evidence of a potential trigger of kidney protein degradation, we investigated, by applying an *ex-vivo* model of precision cut human kidney slices and an *in-vitro* model of immortalized human kidney podocytes, whether TGF-β-Calpain interaction may drive degradation of key structural proteins destabilizing the podocyte cytoskeleton.

## 2. MATERIALS AND METHODS

### 2.1 Clinical characteristics of donor cohorts

This study is based on the analysis of pre-transplantation kidney biopsies obtained from 65 deceased and living kidney donors. Deceased donor kidney biopsies were obtained from the Quality in Organ Donor (QUOD) biobank, a national multicenter UK wide bioresource of deceased donor clinical samples procured during donor management and organ procurement^25^. Healthy living donor (LD) kidney biopsies were collected by the Oxford Transplant Biobank (OTB).

Selection of donor kidneys was based on paired 12month post-transplant outcomes. To minimize the impact of recipient factors on outcomes we only included kidneys for which the contralateral kidney was transplanted and had similar 12-month post-transplant outcome; either suboptimal outcome (SO) or good outcome (GO). For DBD, average 12-months post-transplant estimated (e)GFR of suboptimal outcome (SO) grafts was 31 ± 9 ml/min/1.73 m^2^ (mean (±SD), and for good outcome (GO) was 82 ± 22 mL/min (mean (±SD) per 1.73 m^2^). For DCD, average 12-months post-transplant eGFR of SO grafts was 29±9 ml/min/1.73mm^3^ and for GO grafts was 77±17 ml/min/1.73mm^3^.

All clinical samples were linked to corresponding donor and recipient demographic and clinical meta data, provided by NHS Blood and Transplant registry (NHSBT) as described in Table 1. Biopsies were selected from one kidney per donor, either the left or right kidney, selected at random. The biopsies were then divided in two, with one half stored in RNAlater, then liquid nitrogen and the other stored in formalin.

**Data Table 1.**
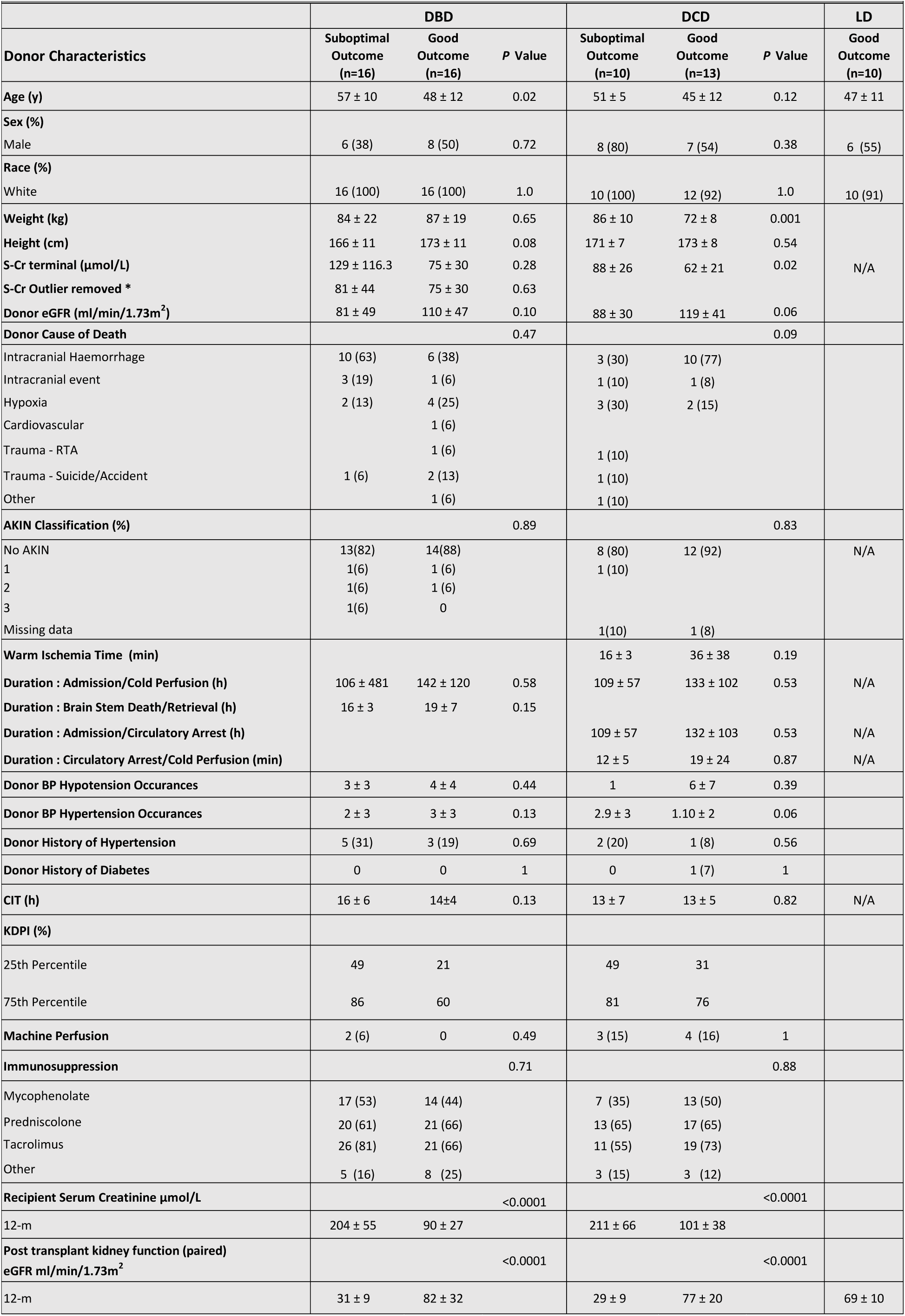
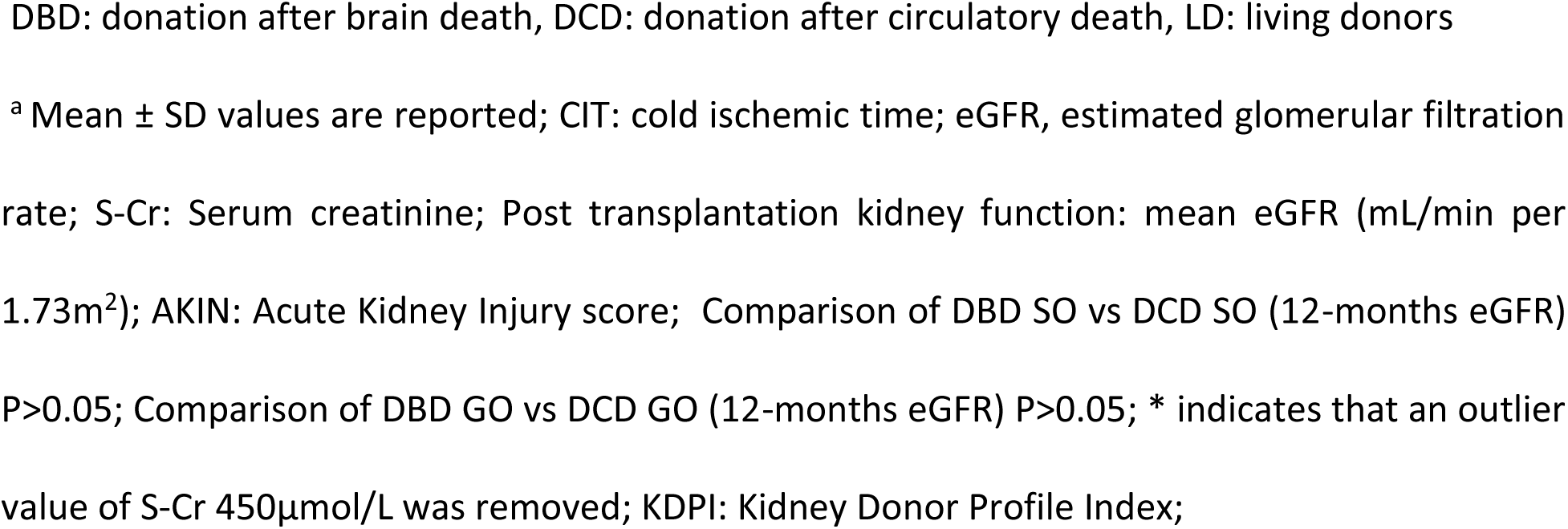
Donor and recipient charactristics

Deceased and living donor biopsies were collected according to the same stringent predefined collection protocols. Deceased and living donor kidney biopsies were collected *ex situ* from the upper pole of the donor kidney cortex during back table preparation in the donor hospital, using a 23mm needle biopsy gun. All donor (deceased and living) kidney biopsies were procured with identical protocols to minimize pre-analytical variability and technical precautions were taken to eliminate ex-vivo sample proteolysis.

For the initial degradomics profiling by PROTOMAP, we analysed two cohorts of pooled biopsy homogenates of DBD (n=10) kidneys with contrasting transplant outcomes SO vs GO (Figure 1A&B). Degradomics data were confirmed by Western blot on an independent cohort of DBD (n=22) donor kidneys. Biopsies from DCD donor kidneys (n=23) with suboptimal outcomes (SO), good outcomes (GO) and from living donors (LD, n=10) with good 12-month transplant outcomes were included as a reference cohort. Donor and recipient characteristics are shown in Table 1.

**Figure 1.**
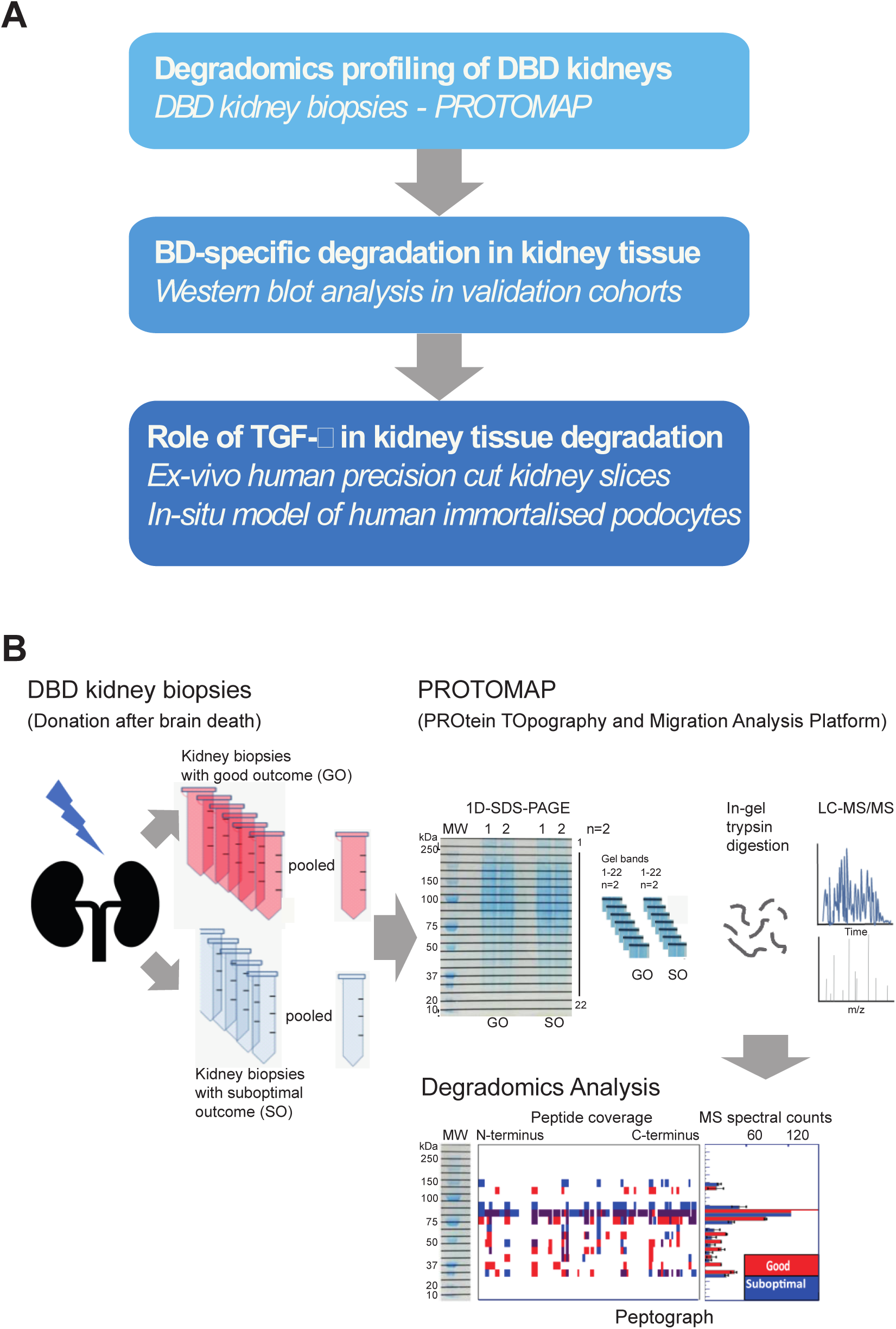
Experimental and technical workflows used in this study. ***A.*** *Study design.* We first studied the degradation profiles of DBD kidneys that had contrasting 12-month post-transplant outcomes; either suboptimal (SO) or good allograft (GO) function. We applied the PROtein TOpography and Migration Analysis Platform (PROTOMAP) in kidney biopsies that were obtained from the QUOD biobank. The analysis shortlisted 8 podocyte proteins that were proteolytically cleaved in SO DBD kidneys. Western blotting on a validation cohort of deceased (DBD& DCD) and living donor (LD) kidney biopsies showed brain death specific degradation patterns. To explain the degradation profiles, we tested the role of TGF-β in kidney tissue degradation by employing an *ex vivo* model of precision cut human kidney slices and an *in vivo* model of human podocyte cells; ***B.*** *Description of the PROtein TOpography Migration Analysis Platform (PROTOMAP) workflow.* DBD kidney biopsy protein homogenates were first separated by SDS- electrophoresis, divided in to 24 horizontal sections and subsequently analysed by LC-MS/MS. Bioinformatics analysis combined data from the gel electrophoresis and mass spectrometry spectra to generate peptographs that provided information on degradation profiles of the intact protein and the generated fragments. The resulting peptographs were screened to identify proteins with evidence of endogenous proteolysis and generation of fragment intermediates at a lower molecular weight than the full length. The selection of proteins for further analysis was based on the following factors: i) increased spectral counts of fragments that had migrated at a lower molecular mass in SO (blue bars) versus GO (red bars), ii) enrichment of the intact protein or protein fragments in the SO group and iii) biological relevance, with an emphasis on cytoskeletal proteins involved in the podocyte.

### 2.2 Study Approval and Ethics statement

Informed consent from donor families was obtained prior to sample procurement. Collection of QUOD samples and the research ethics approval was provided by QUOD (NW/18/0187). The collection of living donor samples and research ethics approval was provided by Oxford Transplant Biobank (OTB) 19/SC/0529.

### 2.3 Sample preparation

RNA later treated kidney biopsies were homogenised in RIPA buffer (89900, Thermo Scientific, Illinois, USA) with protease and phosphatase inhibitor cocktail (1861280, Thermo Scientific, Illinois, USA) using a bead beater (Biorad, Hertfordshire, UK) at 6500rpm for three cycles of 40 seconds with intermediate cooling on wet ice between cycles. Biopsy protein concentration was determined using a Pierce Bicinchoninic Acid protein assay kit (23227 Thermo Scientific, Illinois, USA). Remaining lysates were aliquoted and stored at -80°C until analysis.

### 2.4 Protein Topography and Migration analysis platform (PROTOMAP)

Kidney biopsies from the QUOD biobank were prepared and analysed by PROTOMAP as previously described^22, 26, 27^. In brief, PROTOMAP analysis was used to compare the profiles between two experimental cohorts; suboptimal (SO) and good outcomes (GO). Five biopsies per clinical outcome (SO vs GO, n=5 each) were pooled to a final aliquot of 100μg. Pools were denatured and reduced in standard Laemmli buffer with DTT and divided into two equal aliquots and separated by SDS-PAGE (NOVEX, Invitrogen 4-12% gradient, Thermo Fisher Scientific). The gels were stained with Coomassie instant blue and each pooled sample lane cut into 22 horizontal slices, generating 44 samples overall. The gel pieces were subjected to in-gel trypsin digestion as previously described^27^. Briefly, gel pieces were de-stained in a solution of 1 ml 50 % methanol, 5 % acetic acid in Milli-Q-H2O solution until transparent, then dehydrated using 200 µl acetonitrile (ACN) for 5 min. Proteins in gel pieces were reduced by addition of 30 µl of 10 mM dithiothreitol (DTT) for 30 min followed by alkylation with 30 µl of 50 mM iodoacetamide (IAA) for 30 min. Gel pieces were dehydrated with 200 µl ACN, resuspended in 30 µl 100 mM ammonium bi-carbonate containing 20 ng/µl trypsin and incubated overnight at 37 °C with gentle mixing. Peptide digests were extracted from the gel matrix using 50 µl extraction buffer I (50 % ACN, 5 % FA) followed by 50 µl extraction buffer II (85 % ACN, 5 % FA), collected and dried by vacuum centrifugation. Pellets were resuspended in 30 µl of buffer A (98 % Milli-Q-H2O, 2 % acetonitrile, 0.1 % formic acid) and analysed by nano ultra-high performance liquid chromatography tandem mass spectrometry (LC-MS/MS) as described previously ^28^.

### 2.5 Analysis of PROTOMAP derived mass spectrometry data

The PROTOMAP method incorporates the protein migration patterns on SDS-PAGE electrophoresis with peptide sequence coverage and spectral counts acquired by LC-MS/MS analysis, allowing the results to be visualised as peptographs for every identified protein^26, 27, 29^ To do this, the raw MS data was converted to Mascot generic files using msconvert (Proteowizard, version 2) and further analysed as described^27, 30, 31^. Briefly, MS/MS spectra data were searched using Mascot v2.5.1 against the UniProt *Homo sapiens* reference proteome (version 2017). Mascot results were then exported as DTASelect files with a false discovery rate threshold of 1% and analysed using the PROTMAP perl scripts obtained from http://www.scripps.edu/cravatt/protomap/. The data were then processed generating the peptographs using custom Perl scripts as previously described^22^.

Each of the peptographs (Figure 1B) has two panels. The left shows the protein sequence overage from N- to C-terminus in each band. Peptide sequences represented in red are from GO donors and in blue from SO donors; whilst the purple fragments are common to both experimental cohorts. The right panel shows the relative quantitation using spectral counts for each protein between the two pools. Protein degradation was defined by fragments with spectral counts detected at a lower molecular weight compared to the expected size of the full-length protein. As the analysis included technical duplicates for each sample, for each resulting peptograph, spectral counts were averaged for each band and condition and displayed with error bars representing standard errors of the mean.

#### Pathway analysis

Stringent selection criteria were applied for the analysis and shortlist of protein peptographs with the requirement for at least 20% protein sequence coverage for shortlisting proteins. The selected dataset of 2,320 protein IDs and 1554 protein classes was entered to PANTHER classification system^32^ v 14.0 (http://pantherdb.org/) to cluster the proteins according to biological function. Protein datasets of specific clusters were further analysed to identify protein - protein interactions, prevalent KEGG pathways and pathway enrichment using STRING^33^ version 11.0, (https://string-db.org/).

The mass spectrometry proteomics data have been deposited to the ProteomeXchange Consortium via the PRIDE partner repository with the dataset identifier PXD022074 and 10.6019/PXD022074.

### 2.6 Precision cut human kidney slices

#### Ethics statement

The use of human tissue for the preparation of Precision Cut Kidney Slices (PCKS) was approved by the Central Denmark Region Committees on Biomedical Research Ethics (Journal number 1-10-72-211-17) and The Danish Data Protection Agency. All participants gave written informed consent.

### 2.7 Preparation of PCKS

PCKS were prepared from functional (*i.e.* eGFR >60 ml/min/1.73m²) and macroscopically healthy renal cortical tissue obtained from patients following tumor nephrectomies, as described previously^34^. In short, slices were prepared in ice-cold Krebs-Henseleit buffer and saturated with carbogen (95% O_2_, 5% CO_2_), using a Krumdieck tissue slicer. Subsequently, PCKS were cultured in William’s E medium with GlutaMAX containing ciprofloxacin and D-(+)-Glucose solution at 37°C in an 80% O_2_, 5% CO_2_ atmosphere while gently shaken. PCKS viability was assessed by determining the ATP content of the slices using the ATP Colorimetric/Fluorometric Assay Kit (Sigma-Aldrich, Søborg, Denmark), according to the manufacturer’s instructions.

Human PCKS were treated with TGF-ß (10 ng) (R&D Systems, Denmark) prepared in William’s E medium with GlutaMAX containing ciprofloxacin and D-(+)-Glucose solution for 24 hours at 37°C. Control PCKS were treated with William’s E medium with GlutaMAX containing ciprofloxacin and D-(+)-Glucose solution only, for 24 hours.

### 2.8 Cell Culture

Conditionally immortalised human podocyte cell lines isolated from donated kidneys, as previously described ^35^ were provided by the Bristol Renal Group at the University of Bristol (Bristol, UK). The podocytes were cultured in RPMI1650 (Sigma, UK) supplemented with 10% fetal calf serum (FCS) at 33°C to encourage proliferation. After growing to 70% confluence, cells were treated with trypsin and re-seeded into 6-well plates and incubated at 33°C. Once the cells had grown to 70% confluence, they were transferred to 37°C for 10 days.

#### TGF-ß Treatment

Podocytes were treated with varying concentrations of TGF-ß (5-20ng) (R&D Systems, UK) prepared in 4mM HCL with 1mg/mL BSA added to RPMI1650 for up to 24hours at 37°C. Control cells were treated with RPMI1650 only, for 24hours.

#### Calpeptin Treatment

In inhibition experiments podocytes were treated with the Calpain-1 inhibitor, Calpeptin (Sigma, Dorset, UK). Cells to be treated with Calpeptin were pre-treated with the inhibitor for 3 hours prior to treatment with TGF-ß. Calpeptin was then applied with TGF-ß treatment also and used at a concentration of 1μM in all experiments.

Following treatment, cells were collected by treating with trypsin for 3 minutes at 37°C followed by gentle scraping. Cells were centrifuged, supernatant removed, and the media replaced with 50 μL of RIPA buffer containing phosphatase and protease inhibitor cocktail as above. Cells were then sonicated using a probe sonicator (Fisher Scientific, Loughborough, UK) for 20 seconds at 15 amps for 2 cycles, samples were cooled on wet ice between cycles. Protein concentration was determined using by BCA and homogenates aliquoted and stored at -80°C.

### 2.9 Western Blotting

Cell and kidney biopsy homogenates containing 1μg/μL of protein were denatured at 70°C in Laemmli buffer and separated using 4-12% pre-cast Bis-Tris gels (Thermo Fisher Scientific, Massachusetts, USA), in MOPS running buffer then transferred onto a PVDF membranes (Thermo Fisher Scientific, Massachusetts, USA); according to standard protocols. Membranes were blocked in PBS (Thermo Fisher Scientific, Massachusetts, USA) with 0.1% Tween-20 (Sigma, Missouri, USA) and milk, then incubated with primary antibodies overnight at 4°C. Antibodies used were; rabbit polyclonal anti-calpain-1 (2556S, Cell Signalling), mouse monoclonal anti α-actinin-4 (SC390205, Santa Cruz Biotechnology), goat polyclonal anti Talin-1 (AF5456, R&D Systems) and rabbit anti GAPDH (5174s, Cell Signalling). Finally, the membranes were incubated in species appropriate secondary antibodies according to manufacturer’s recommendations (anti-rabbit IgG (7074, Cell Signalling, London, UK) anti-mouse IgG (7076, Cell Signalling, London, UK) and anti-goat IgG (HAF109, R&D Systems, Abingdon, UK)) and imaged using the LI-COR Odyssey system (LiCOR, Nebraska, USA). Analysis and quantification were performed using LiCOR Image Studio Lite (Version 5.2). Full length proteins and proteolytic fragment intensities were normalized against GAPDH. To determine proteolytic processing the ratio of fragment to full length intensities was calculated.

### 2.10 Calpain Activity

Calpain activity was measured in differentiated podocyte cells as well as PCKS using a Calpain Activity Assay Kit (ab65308, Abcam, Cambridge, UK), according to the kit protocol and as previously described ^36^. In brief, cells were collected with gentle scraping, centrifuged, supernatant removed and washed in cold PBS. Cells and slices were then lysed in the provided buffer and protein concentration determined by BCA assay kit. Between 40-100μg of cell protein was loaded onto a 96-well plate and incubated with provided buffers and Calpain substrates. Control wells were treated with either active Calpain (positive control) or Calpeptin (negative control). Plates were incubated and absorbance read at 350/535nm.

### 2.11 Cell Imaging

Cells were seeded into a 96 well clear bottom black walled plate and differentiated at 37°C for 10 days. If Calpeptin treatment was required, this was applied at 1μM for 3 hours before addition of TGF-ß. Cells were then washed twice with PBS before fixing with 4% paraformaldehyde (PFA) for 20 minutes and permeabilising with 0.1% TritonX100 for 5 minutes. Cells were blocked in 5% BSA overnight. F-actin structures were visualised using Alexa fluor phalloidin 647 (Invitrogen). Nuclear regions were defined by Hoechst 33342 stains. Cell imaging was automated using an IN Cell Analyzer (GE Healthcare) high content imaging platform with a 10x objective. Analysis was performed using the *IN Cell* Analyser developer software. Actin fibers were counted and measured with cells being defined by Hoechst and phalloidin staining. Three technical replicates were performed within each experiment, with 4 fields of view per well; yielding data for around 2000 cells per condition, per experiment.

### 2.12 QPCR

Total RNA was isolated using a NucleoSpin RNA II mini kit (Macherey Nagel), following the manufacturer’s instructions. RNA was quantitated by spectrophotometry and stored at −80°C. cDNA was synthesized from 0.5 μg RNA with the RevertAid First Strand synthesis kit (Thermo Scientific). QPCR was performed using 100 ng cDNA, which served as the template for PCR amplification using the Brilliant SYBR Green qPCR Master Mix (Thermo Scientific), according to the manufacturer’s instructions. Primers were: Collagen 1A1 (NM_000088.3): forward 5′- CCTGGATGCCATCAAAGTCT-3′ and reverse 5′-AATCCATCGGTCATGCTCTC-3′; RPL22 (NM_000983.3): forward 5′-TCGCTCACCTCCCTTTCTAA-3′ and reverse 5′- TCACGGTGATCTTGCTCTTG-3′.

### 2.13 Sirius Red staining and Masson’s Trichrome staining

PCKSs were immersed in 4% PFA for 1 hour, rinsed with PBS, dehydrated using a graded series of alcohol and embedded in paraffin. Tissue sections (5 µM) were deparaffinised and rehydrated. Sirius red sections were left in tap water for 10 min. Followed by 30 min in 0.1 % Picro Sirius solution (Ampliqon). Then sections were dehydrated and mounted with Mounting Medium Pertex (HistoLab). *Masson’s Trichrome staining*. The sections were left in Bouin’s solution overnight and then rinsed in running tab water for 5 minutes in order to remove the picric acid. After rinsing the sections were colored in Mayer’s Haematoxylin for 10 minutes to stain the nucleuses, rinsed in running tab water, and then colored in Biebrich scarlet for 5 minutes. Afterwards, the sections were placed in 5% Phosphomolybdic Acid solution for 10 minutes and then directly into Aniline blue for 5 minutes. Sections were then incubated with 1% Acetic acid and dehydrated through increasing solutions of ethanol before being mounted with cover glasses using Tissue Mount. The two stains were assessed by capturing 5 pictures from each slice at x40 magnification and measuring the area of staining in percentage of total tissue area using ImageJ software (version 2017).

### 2.14 Statistical Analysis

The number of samples required to demonstrate meaningful changes between donor subgroups with extreme graft function either suboptimal or good outcomes has been calculated based on the effect size observed in previous work^23^. Taking into consideration that paired posttransplant outcomes reduces variability we calculated that an estimated minimum of n=8 individual donor samples were needed to achieve P<0.05, a statistical power 0.8% and a difference 0.2% on the expression of previously identified proteins between SO vs GO.

Statistics for this study were performed using Graph Pad Prism V7. Results were expressed as mean ±SD. Differences in continuous variables were analysed using an ANOVA (more than two groups) or two-tailed unpaired Mann-Whitney test whilst discrete variables were analysed using χ^2^ tests. Minimum of three technical replicates were performed in each experimental analysis. Kidney Donor Profile Index was calculated using the on-line calculator https://optn.transplant.hrsa.gov/resources/allocation-calculators/kdpi-calculator

## 3. RESULTS

### 3.1 Clinical characteristics of deceased and living donors

Kidney biopsies were obtained from DBD, DCD and living donors at the back table immediately after kidney procurement. To minimize the impact of cofounding factors related to post-procurement and recipient factors, donors were selected on the basis of paired 12-m post-transplant outcomes. Clinical metadata confirmed that the selected donor groups were representative of the donor population in UK and shown limited clinical variability (Table 1). The DBD SO group with median age (57+/-10y) were older than DCD SO (51+/-5y) but the age difference was not considered to be clinically significant. There was also no correlation of protein fragment intensity with donor age as described in Table 2. Intracranial event (trauma/hemorrhage) was the main cause of death for DBDs and DCDs. There were more DBD donors with AKIN 1-3 than DCD, likely to reflect the uncertainty of clinicians in accepting DCD kidneys with AKIN>1. Duration of donor admission to cold perfusion, WIT and CIT were all comparable among all donor subgroups. KDPI profiles revealed clear differences in the 25^th^ and 75^th^ percentiles between SO and GO in both donor types, notably there was no difference between DBDs and DCDs. In the associated metadata has been recorded that one of the DBD SO donors had a 5.4-fold increase of serum creatinine (from admission to terminal) and this increased the mean S-Cr value of DBD SO to reach significance when compared to DBD GO. If we exclude this outlier value, then the mean of terminal S-Cr DBD SO (81+/-44) is comparable to DCD SO (88+/-26) (µmol/L). Recipients mean 12-month post-transplant eGFR was significant different between SO vs GO (SO vs GO; *P*<0.0001).

**Table 2.**
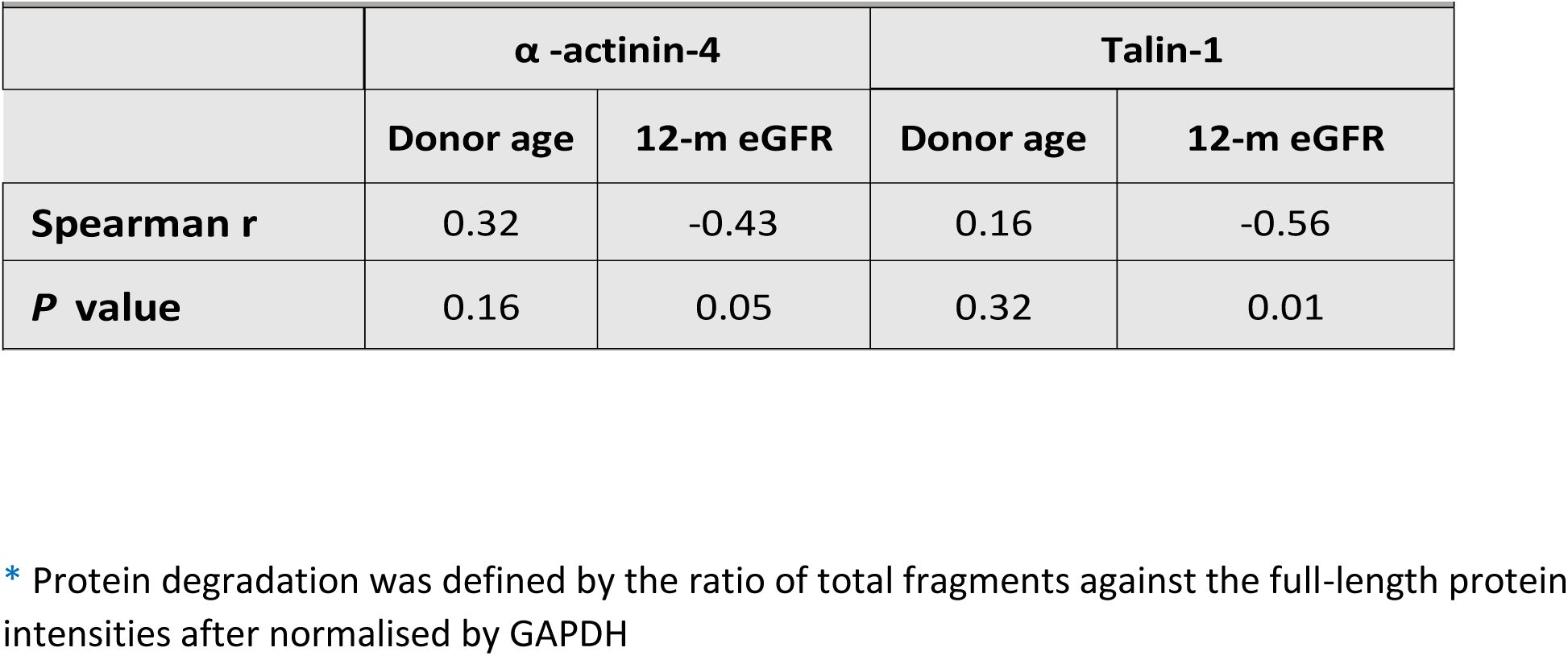
Correlation of protein degradation* to donor age and post-transplant 12-m eGFR

|  |  |  |  |  |
| --- | --- | --- | --- | --- |
|  | **$\alpha$ -actinin-4** |  | **Talin-1** |  |
|  | **Donor age** | **12-m eGFR** | **Donor age** | **12-m eGFR** |
| **Spearman r** | 0.32 | -0.43 | 0.16 | -0.56 |
| **P value** | 0.16 | 0.05 | 0.32 | 0.01 |

\* Protein degradation was defined by the ratio of total fragments against the full-length protein intensities after normalised by GAPDH

### 3.2 PROTOMAP shows degradation alterations associate to extreme transplant outcomes

Putative differences in the pre-implantation kidney degradome of SO and GO DBD kidneys were mapped through the Protein Topography and Migration Analysis Platform (PROTOMAP)^27, 37^ (Figure 1B).

This analysis generated 15,700 peptographs corresponding to 6,700 proteins. Stringent selection criteria (including the requirement for at least 20% protein sequence coverage for shortlisting proteins) reduced the data set to 2,320 protein IDs and 1554 protein classes. Pathway analysis performed using (PANTHER 16.0) mapped 135 of these proteins as cytoskeletal and 65 of these linked to the integrin regulation pathway. Direct protein-protein interactions (STRING 6.0) showed enrichment of KEGG pathways of focal adhesion false discovery rate (FDR) 2.11 e-33, catabolic regulation FDR 1.06 e-09 and integrin mediated signaling FDR 3.09e-9 (Supplementary SF1).

To identify evidence of increased proteolysis in donor kidneys, we scanned the peptographs of the cytoskeletal proteins to detect the generation of lower molecular mass fragments of intact proteins in SO kidneys (blue bars) not observed in GO kidneys (red bars) (Figure 1B). Eight proteins mapped to podocyte cytoskeleton; α-actinin-1, -4, Talin -1, -2, Utrophin, Laminin β2, Synaptopodin, Integrin α-1, showed specific and contrasting degradation profiles for SO when compared to GO grafts (Figure 2, Supplementary SF1 & SF2). To confirm protein degradation detected in PROTOMAP data and further investigate whether there were donor type specific differences in the degradome, we performed a targeted analysis of α-actinin -4 and Talin -1 by Western blot on deceased and living donor kidney biopsies. The cohort of samples selected for blotting was independent of the samples pooled to PROTOMAP analysis.

**Figure 2.**
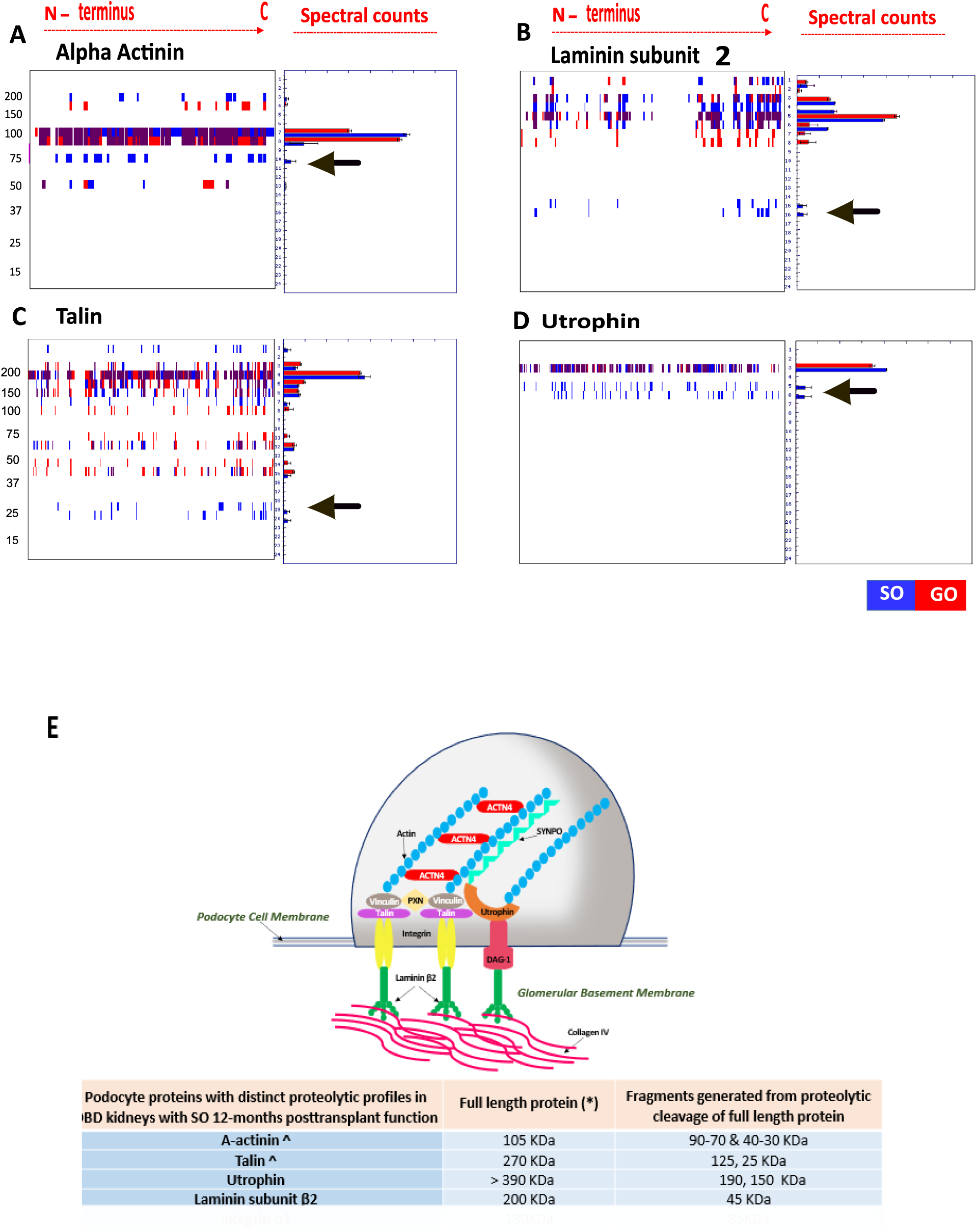
Mapping the generation of protein fragments using PROTOMAP analysis of kidney biopsies. Arrows show the protein fragments detected in lower molecular weight generated by the full-length protein. Blue depicts suboptimal outcome (SO) kidneys and red depicts good outcome (GO) kidneys. Peptographs depict proteins shortlisted from the PROTOMAP analysis of DBD kidneys with GO (**red bars**) pooled n=5; compared to biopsies obtained from DBD kidneys with SO (**blue bars**) pooled n=5. Degradation profiles (noted with the arrow) from N- terminus to C- terminus of α- actinin (A), Laminin β2 (B), Talin (C), Utrophin (D); (**E**) Podocyte schematic showing intracellular actin filaments linked to α-actinin-4 (ACTN4), Synaptopodin (SYNPO), Utrophin, Talin and through Integrins with intertwined Laminin β2 podocyte connected to the podocyte basement membrane (peptographs of Integrin and synaptopodin listed in SF2); (**H**) The table shows the full length and fragments identified from PROTOMAP analysis. As α - actinin-1/-4 isoforms and Talin-1/-2 isoforms share 85% and 76% homology we included both isoforms of each protein in the analysis. * indicates the full-length protein. ^ Indicates that α-actinin and Talin were validated by Western blot on an independent cohort of biopsies and the association of the degradomics profiles to protein isoforms of α-actinin-4 and Talin-1 were confirmed. (The sperate protein isoform peptographs of α-actinin and Talin in addition to the rest of the peptographs are displayed in Supplementary Fig. 2).

### 3.3 Proteolytic degradation profiles were increased in DBD kidneys with suboptimal 12- months post-transplant function

Western blots of α-actinin -4 and Talin -1 of tissue homogenates from SO and GO DBD kidneys (n=32) confirmed the degradome findings (Figure 3 & 4). Correlation analysis excluded associations of kidney tissue protein degradation fragments with donor age but confirmed significant association to 12-month eGFR (Table 2). To explore whether the degradation signatures were altered in the different donor types, we also profiled kidney biopsies from SO and GO DCDs (n=23), and from LDs (n=10).

**Figure 3.**
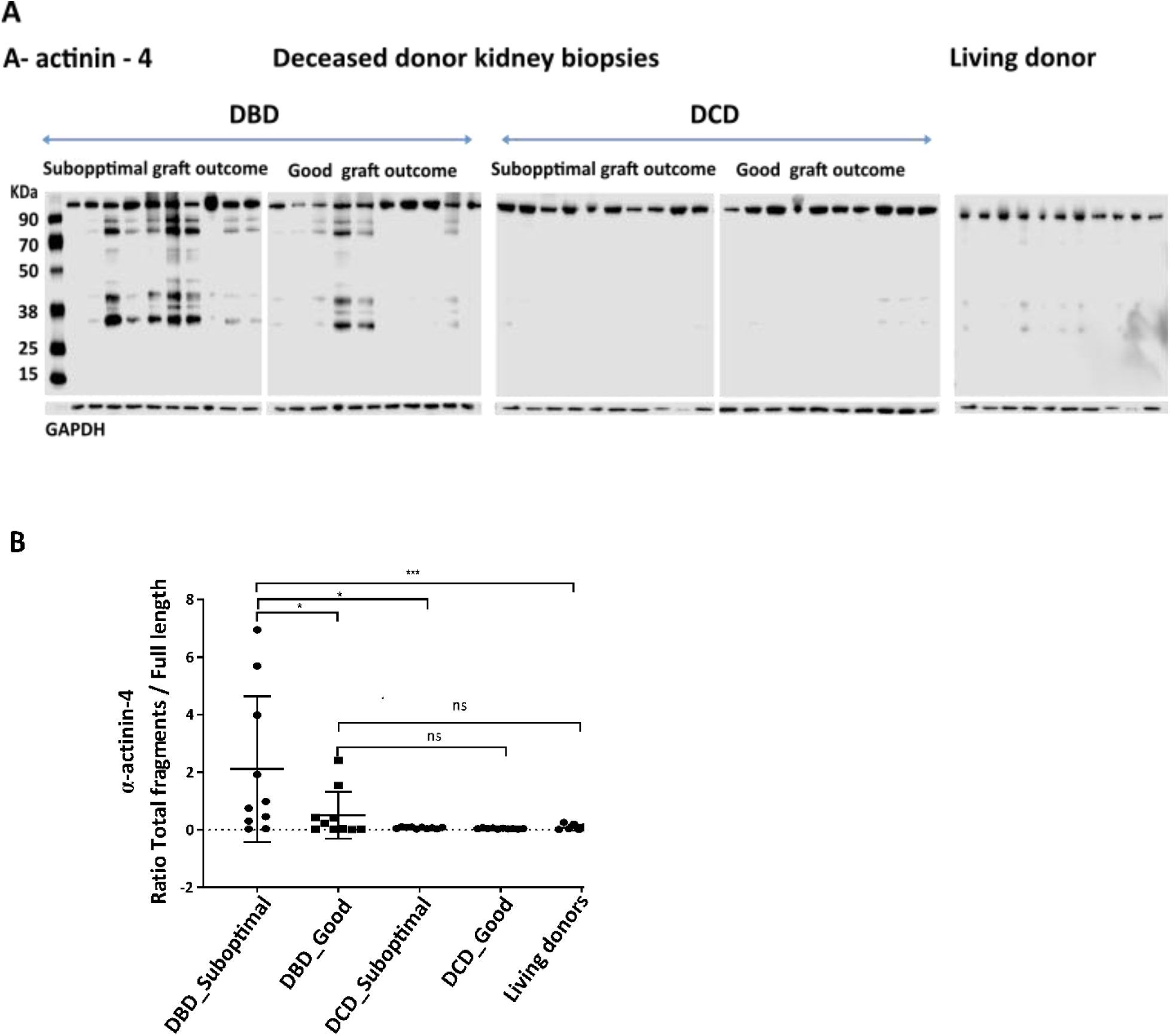
α -actinin-4 proteolytic profile is specific to DBD kidneys and associates to post-transplant outcome. **A.** Western blot analysis of donor kidney biopsies obtained from n=10 DBD with suboptimal outcome (SO: 12-months eGFR (±SD) 31 ± 9 ml/min/1.73 m^2^), n= 10 DBD with good outcome (GO:12-months mean eGFR (±SD) 82 ± 22 mL/min per 1.73 m^2^), n=10 DCD with SO (12-months mean eGFR (±SD) 29 ±9 ml/min/1.73 m2 n=10 DCD with GO 12-months mean eGFR (±SD) 77 ± 17 mL/min per 1.73 m2, and biopsies from n=10 LD shows that the degradation of a-actinin-4 is specific to DBD kidneys while DCD and LD kidneys show no evidence of degradation. α-actinin- 4 shows a distinct profile of fragments generated between 90-70 KDa and between 40-30KDa; **B.** The ratio of total fragment intensities (normalised to GAPDH) to the full-length protein (100 KDa) shows that the proteolytic processing of the DBD kidneys with SO is significantly different to DBD GO * P<0.05) the DBD SO is significantly different to DCD SO and LD (* P<0.05; *** P<0.001) while there was no difference in the degradation pattern between DCD and LD.

**Figure 4.**
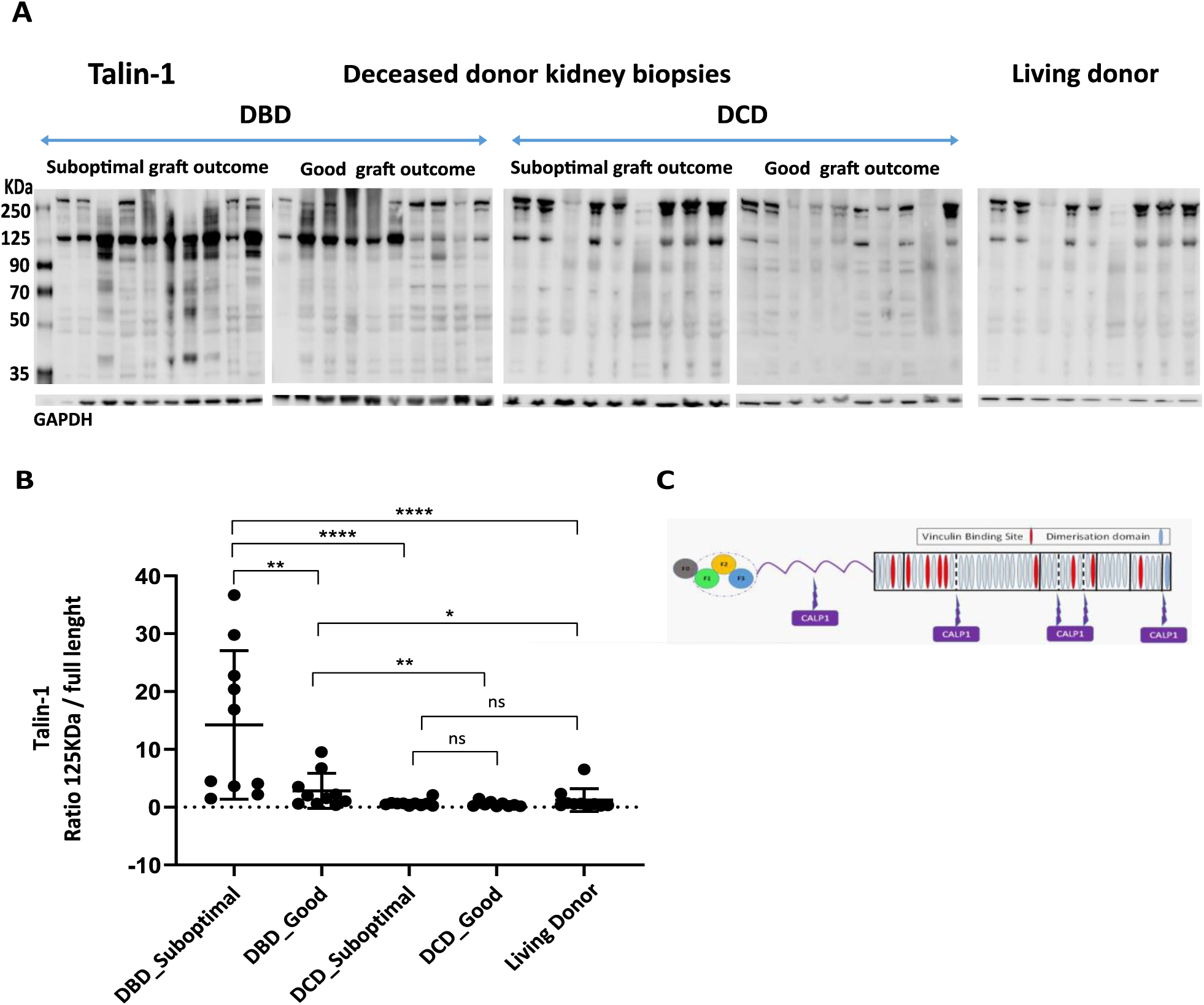
Talin-1 proteolytic profile is specific to DBD kidneys and associates to post-transplant outcome. **A.** Western blot of donor kidney biopsies obtained from n=10 DBD with Suboptimal Outcome (SO: 12-month eGFR (±SD) 31 ± 9 ml/min/1.73 m^2^), n= 10 DBD with Good Outcome (GO: 12- month mean eGFR (±SD) 82 ± 22 mL/min per 1.73 m^2^), n=10 DCD with SO (12-month mean eGFR (±SD) 29 ±9 ml/min/1.73 m2 n=10 DCD with GO 12-month mean eGFR (±SD) 77 ± 17 mL/min per 1.73 m2, and biopsies from n=10 LD shows that the degradation of Talin is specific to DBD kidneys. A fragment at 125kDa is clearly increased with the degradation of the full length Talin protein at 270kDa in the DBD kidneys. An additional fragment at 100kDa was also observed in the DBD SO kidney. The 125kDa fragment was detected in the DCD and LD kidneys also however in both donor groups the full-length protein was clearly distinct; **B.** The ratio of total fragment intensities (normalised to GAPDH) to the full-length protein (270kDa) shows that the proteolytic processing of the DBD kidneys with SO is significantly different to DBD GO (** *P*<0.01), the DBD SO is significantly different to DCD SO and LD (**** *P*<0.0001) while there was no difference in the degradation rates between DCD and LD; **C.** Talin consists of an N-terminal head and a flexible rod. The head and rod are joined by a linker region that is cleaved by Calpain.

First, we examined the peptidomic profile of α-actinin-4 due to its pivotal role in maintaining healthy podocyte function^19, 38^. The profile of α-actinin-4 showed clearly that protein degradation had occurred only in DBD kidneys but not in DCD or LD kidneys (Figure 3A). The detected fragments generated from the full-length 105KDa protein were significantly enhanced in SO, compared to GO DBD kidneys (P<0.05), to DCD (P<0.05) and LDs (P<0.001) (Figure 3B).

Next, we surveyed the degradome profile of Talin-1 due its key structural role in linking integrins to the actin cytoskeleton sustaining the integrity of the podocyte filtration barrier and podocyte membrane function ^39, 40^. The peptidomic profile of Talin demonstrated that in DBDs the full-length 270KDa Talin protein was proteolytically processed with the loss of intact protein (Figure 4A) with a significant relative increase of ∼100-125KDa fragments, observed in all samples, seen in SO DBDs compared to GO DBDs, DCDs and LD (Figure 4B).

Talin is a specific target protein for the non-lysosomal cysteine protease Calpain^41^ with at least 4 calpain cleavage sites (Figure 4C). To further investigate proteolytic cleavage of Talin-1 we investigated Calpain activation in DBD kidneys with SO outcomes.

### 3.4 Peptidomic alterations indicate Calpain-1 activation

Our PROTOMAP data revealed that Calpain-1 had a distinct fragmentation profile in SO kidneys compared to GO-DBD (Figure 5A). This was confirmed by Western blot analysis on a separate sample cohort (Figure 5B) that showed that observed fragment intermediates at ∼50, 40, & 30kDa were significantly increased in SO when compared to GO DBDs (P<0.05) (Figure 5C-E). A distinct fragment ∼18KDa was only seen in SO DBD kidneys (Figure 5F). The Calpain-1 fragments (50, 40, 30, 18KDa) seen enriched in SO DBDs have been previously reported to be associated to Calpain-1 enzymatic activation and subsequent autolytic processing^42, 43^. As we were unable to perform Calpain activity assays in tissue homogenates^44^, we used an *ex vivo* model of precision cut human kidney slices and an *in vitro* model of immortalised human kidney podocyte cells to first validate our findings and then to explore a potential causal pathway to explain the degradomics patterns in DBD kidney biopsies.

**Figure 5.**
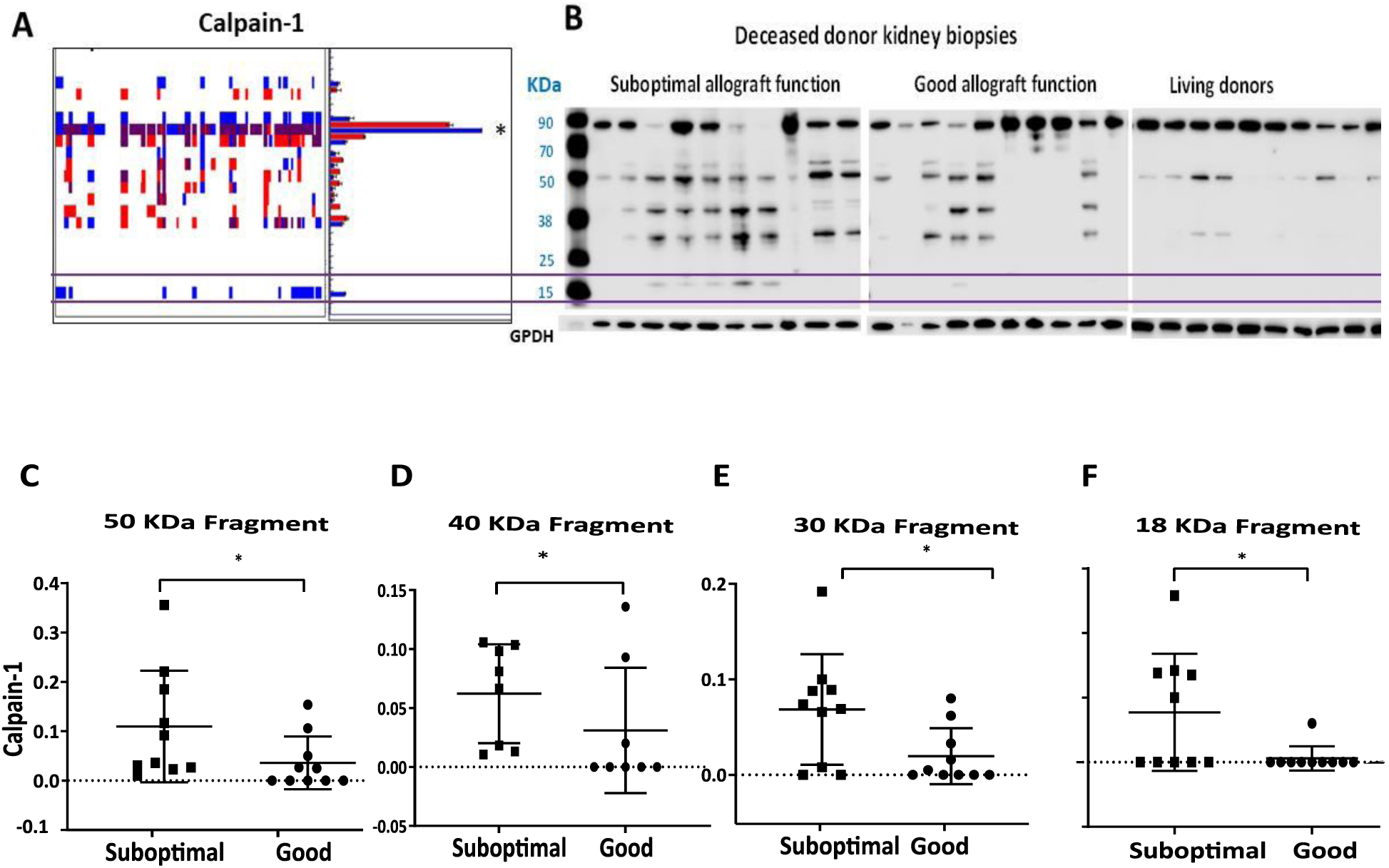
Calpain-1 peptidomic profile reveals enhanced enzymatic activation in suboptimal outcome DBD kidneys. **A.** PROTOMAP analysis showed the generation of a number of fragments in DBD suboptimal outcome (SO) donor groups and the generation of a unique fragment at around 18 KDa in SO donor kidney biopsies; **B.** Western blot analysis of donor kidney biopsies obtained from n=10 DBD with SO (12-months eGFR (±SD) 31 ± 9 ml/min/1.73 m^2^), n= 10 DBD with good outcome (GO; 12-months mean eGFR (±SD) 82 ± 22 mL/min per 1.73 m^2^) showed a distinct peptidomic profiles that included the generation of protein fragments increased at around ∼50kDa (C), ∼40kDa (D), ∼30kDa (E) and of a unique 18KDa fragment (F) in SO when compared to GO DBD kidneys (*P<0.05; t-test). Band intensities were normalised against GAPDH.

### 3.5 TGF-β treatment of precision cut human kidney slices (PCKSs) increases Calpain protease activity

Brain death related upregulation of Transforming Growth Factor Beta (TGF-β) plays a key role in pre-implantation podocyte injury^45^. Given that we have previously shown increased levels of TGF-β in pre-implantation DBD kidney biopsies with SO post-transplant outcomes ^23^, we hypothesized that TGF-β might trigger Calpain activation. To investigate this, we used a novel *ex vivo* model of precision cut human kidney slices (PCKSs). Indeed, treatment of kidney slices with TGF-β significantly increased Calpain activity of PCKSs (P<0.01) compared to the untreated control in the first 24h of treatment. The treated slices also shown morphological alterations characteristic of fibrotic histological changes that were not observed in the control (Figure 6A-D). Sirius Red and Masson Trichrome stained sections revealed interstitial collagen deposition and increased tubulointerstitial fibrosis of PCKSs treated with TGF-β for up to 24h (Figure 6C). Increased levels of Collagen1A1 mRNA expression were also detected in treated compared to control kidney slices (Supplementary fig.3). To determine whether TGF-β linked Calpain-1 activation has a downstream impact on the integrity of the podocyte cytoskeleton, we employed an *in vitro* model of conditionally immortalised human kidney podocyte cells.

**Figure 6.**
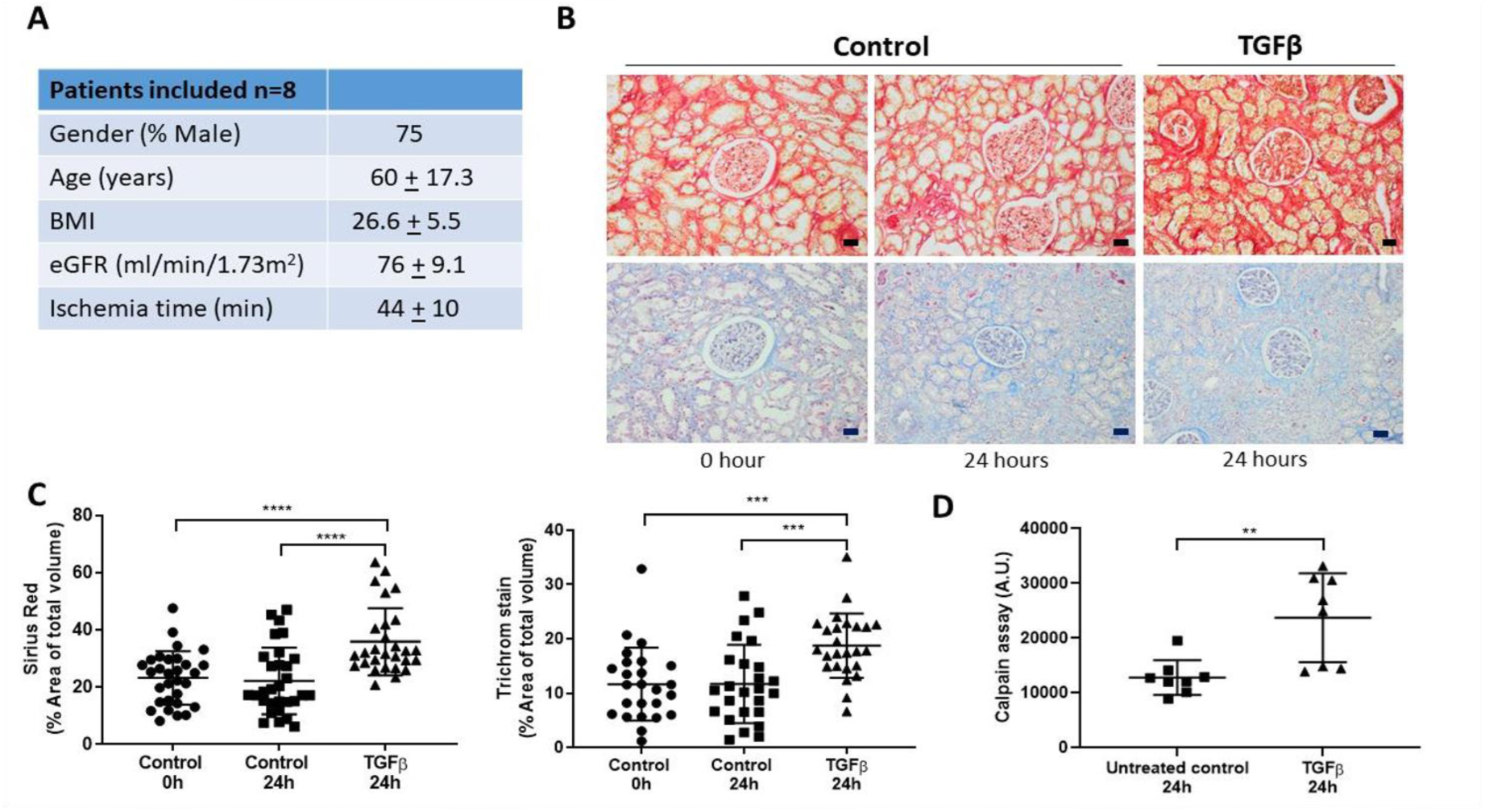
TGF-β t atm nt of h man p*recision-cut kidney slice (PCKS)* increases Calpain activity and caused pro-fibrotic morphological changes. **A.** Table showing the demographic and clinical characteristics of patients who have undergone nephrectomies from where PCKS were prepared, n=8 patients; **B.** PCKS stimulation with TGF-β (10ng) up to 24h caused the kidney slices to develop profibrotic morphological changes detected by serius red (top) and trichrome mason staining (bottom). 20x magnification and 20µm scale bar; **C.** Sirius red and trichrome mason staining revealed that the PCKSs developed profibrotic histological characteristics of cortex, podocyte and tubular structures *(****P<0.0001,***P<0.001; paired t-test; dot plot shows biological and technical replicates)* following treatment with TGF-β up to 24h, (evidence of profibrotic characteristics are derived from Collagen1A1 mRNA comparison as shown in the supplementary data figure 3; measurement of ATP levels showed that the integrity of the PCKS was maintained throughout (Supplementary Fig. 3); **D.** Calpain-1 activity significantly *(**P<0.01; paired t-test)* increased in PCKSs treated with TGF-β up to 24 hours.

### 3.6 TGF-β treatment of conditionally immortalized human kidney podocytes increases Calpain-1 protease activity with subsequent degradation of cytoskeletal Talin-1

We investigated the impact of TGF-β stimulation of human podocyte cells by treating of podocytes with TGF-β for up to 24h. Induced cytoskeletal changes manifested as pro-fibrotic morphological alterations including cell elongation and thickening of the peritubular area (Figure 7A). Importantly, stimulation of podocytes with TGF-β caused significant Calpain activation (P<0.0001) (Figure 7B). Treatment of podocyte cells with Calpain specific inhibitor, Calpeptin (1µM), prior to and during stimulation with TGF-β, prevented TGF-β induced Calpain activation (Figure 7B).

**Figure 7.**
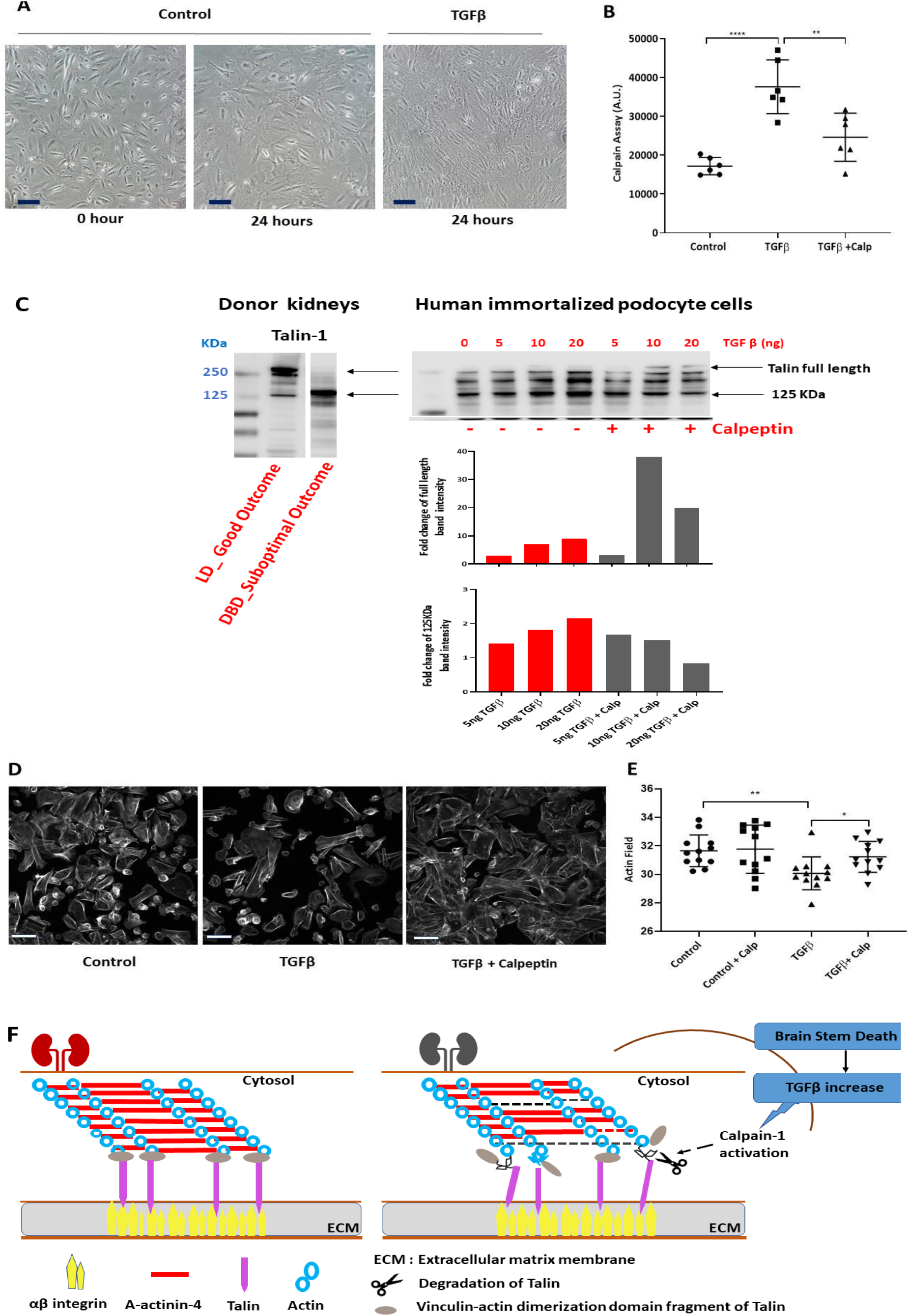
TGF-β t atm nt of h man immo talis d podocyt c lls t i s Calpain activation, actin cytoskeleton dysregulation and cytoskeletal Talin degradation. **A.** Representative contrast microscopy images showing changes to podocyte cells morphology when treated by TGF-β (10ng) for up to 24h; 20x magnification, scale bar is 100µm; **B.** Calpain activity of podocyte cells was significantly increased following stimulation with TGF-β (10ng) up to 24h when compared to control *(****P<0.0001; t-test).* Treatment of cells with Calpeptin (1µM) prior to and during TGF-β stimulation significantly reduced Calpain activation *(**P<0.01; t-test);* **C.** The degradation pattern of Talin-1 of suboptimal DBD kidneys compared to LD kidney with good outcome showed the generation of ∼125KDa fragment (cropped western blot presenting donor kidneys probed with anti-Talin-1 was derived from data presented in Figure 4A). *In vitro* human kidney podocyte cells stimulated with TGF-β (0-20ng, up to 24h) showed a dose responsive loss of the Talin full length protein. Talin-1 proteolytic fragments were detected as ∼190 and ∼125kDa bands with a further lower intensity band at ∼100 kDa. The generation of these fragments was reduced when podocytes were treated with Calpain inhibitor Calpeptin (1µM) prior to and during TGF-β treatment. The two histograms show the alterations (fold changes) in the band intensities of the Talin-1 main band and the 125KDa fragment following treatment of podocytes with TGF-β in the absence (red bars) and presence of Calpain-1 inhibitor Calpeptin (grey bars); **D.** Phalloidin staining of actin stress fibres in cultured conditionally immortalised podocytes in response to TGF-β (10ng) and Calpeptin (1µM) treatments (three technical replicates); **E.** TGF-β treatment (10ng) decreased the length of actin fibres *(**P<0.01; t-test)* indicating dysregulation of the actin cytoskeleton. This loss of stress fibre area was recovered by treating cells with 1µM Calpeptin alongside the TGF-β treatment *(*P<0.05; t-test).* 10x magnification and 20µm scale bar; **F.** Schematic representation of the podocyte cytoskeletal matrix of healthy and after brain death kidney; brain stem death increases TGF-β triggering the activation of Calpain-1 and subsequent degradation of cytoskeletal Talin-1 affecting the integrity of actin cytoskeleton in the kidneys with suboptimal post-transplant function.

Given the key role of Talin-1 in sustaining the podocyte integrity and since we had shown that Talin-1 was markedly degraded in DBD SO kidneys, we examined whether the interaction of TGF-β and Calpain-1 led to degradation of Talin cytoskeletal protein in this *in vitro* model.

Analysis by Western blot of TGF-β treated podocyte homogenates revealed that Talin-1 was proteolytically processed with the generation of distinct fragments at ∼190, ∼125 and ∼100 kDa (Fig. 7C). Notably, the ∼125kDa fragment we had previously shown to be distinct in the DBD SO kidney biopsies with corresponding degradation of the full-length protein (Figure 4A & 7C), was generated in podocytes in a TGF-β concentration dependent manner. Inhibition with Calpeptin prior and during TGF-β stimulation, reduced the proteolytic processing, limited the generation of the 125KDa fragment and reduced cleavage of full-length Talin-1 (Figure 7C). This result strongly suggested that Talin-1 degradation in this model was mediated by Calpain, activated in turn by the treatment of podocyte cells with TGF-β (Figure 7C).

As podocyte motility is a key measurement of podocyte integrity in kidney disease; we examined podocyte motility by determining how TGF-β treatment impacted actin remodeling. Following TGF-β stimulation of podocyte cells, there was significant reduction of the actin stress fibers compared to the untreated cells (P<0.01) and TGF-β+calpeptin treated cells (<0.05) as measured by rhodamine phalloidin staining (Figure 7D & E). Taking together, these data show that TGF-β triggered Calpain activation negatively impacted the integrity of the podocyte actin cytoskeleton. The observed impact was inhibited by Calpain inhibition, preserving the podocyte cytoskeleton.

## 4. DISCUSSION

Our data, obtained from a unique cohort of pre transplant biopsies from DBD, DCD and LD kidneys, provide initial evidence that the activation of proteolytic processes in DBD kidneys causes cytoskeletal alterations that may deem these grafts susceptible to subsequent injury, impacting on post-transplant graft function. Replicating the observed DBD kidney key degradation profiles of Talin-1 in human kidney immortalised podocyte cells treated with TGF-β suggests the podocyte cytoskeleton as a target of proteolytic modifications that merits further investigation.

Brain death donor kidneys are the preferred source of deceased donor transplants worldwide ^8^. While improvements in organ procurement, immune suppression regimes and treatment of postoperative complications have greatly improved transplant outcomes, biological processes related to brain death still exert a detrimental effect on short- and long- term allograft function compared to living donor transplants ^7, 46^. Impaired long-term outcomes result in graft failure, return to dialysis and re-transplantation^4^. As we increasingly accept older and higher risk organs from DBD donors, a better insight into processes associated with brain death impact on donor kidney quality is crucial to reduce injury and design novel targeted interventions to recondition these organs.

To investigate the donor kidney degradome we analysed deceased and living donor kidney biopsies obtained with identical procurement protocols to minimize pre-analytical confounding factors. The selection of donor kidneys was based on paired transplant outcomes that limited the impact of surgical and recipient heterogeneity on post-transplant function. This stringent experimental selection also increased our confidence in obtaining meaningful data with the analysed sample size. Initial degradomics profiling of DBD kidneys with extreme outcomes followed by western blotting of an independent cohort of DBD, DCD and LD kidney biopsies.

We, and more recently others, have shown that the application of the PROTOMAP technique to complex clinical samples provides a comprehensive profiling of both N- and C- terminal proteoforms with sufficient sequence coverage to determine epitope coverage by commercial antibodies, thereby allowing validation by immunoblotting.^27, 47, 48^. Initial, unbiased PROTOMAP degradation profiling and pathway analysis mapped α -actinin, Synaptopodin, Talin, Laminin β2, Integrin α1 and Utrophin as a cluster of cytoskeletal proteins associated with focal adhesion in podocytes had undergone enhanced proteolytic processing in DBD kidneys with SO post-transplant function. Evidence that proteolytic cleavage of key podocyte proteins is a destabilizing process that damages the integrity of podocytes and the glomerular filtration barrier has been previously shown in experimental models but with limited evidence on human kidneys to date^21, 49^. Therefore, to further investigate protein degradation of podocyte cytoskeleton and to better visualize donor specific degradation patterns, we examined degradome profiles in DBD, DCD and LD kidney biopsies by Western blotting on a separate cohort of donor kidney biopsies. Blotting confirmed our PROTOMAP data showing that DBD-specific α -actinin-4 and Talin-1 peptide fragments were enriched in suboptimal outcome grafts indicating that these proteins had undergone proteolytic processing.

Although these two actin-binding proteins had been previously identified, mainly in-vitro and preclinical settings, as susceptible to proteolysis^19, 39^, here we showed that for these proteins, preferential proteolytic modification in DBD kidneys may cause subclinical changes in the grafts leading to post-transplant dysfunction.

We looked in detail the donor and recipient clinical metadata and no clinically associated differences could explain the observed kidney proteolytic profiles between DBD, DCD and LD as there was limited heterogeneity between the subgroups. The DBD-SO group was older than the rest of donor groups, but the mean age was representative of the deceased donor population in UK. Furthermore, we confirmed that there was no correlation of proteolytic fragment generation with donor age. Donor eGFR and AKIN were comparable between donors. Finally, there was a significant negative correlation (p<0.05) of protein degradation fragment generation to graft function as expressed by recipient 12-month eGFR.

We observed that in GO & SO DCDs and LDs the ∼ 125kDa Talin-1 protein fragment co-existed with the Talin-1 full length protein that appeared intact in most samples of these donor cohorts. We suggest that the generation of the ∼125kDa fragment may also results from normal protein turnover as part of the physiological maintenance of cellular homeostasis. In contrast, for the majority of SO DBD kidneys there was a specific loss of Talin-1 full length protein with a significant increase in the ratio of ∼125kDa to full length protein compared to GO DBDs, DCDs and LDs that suggests DBD specific activation of proteolytic processes were increased in SO kidneys. Talin-1 has a pivotal role in integrin activation but also, notably, in linking integrins to actin cytoskeleton to sustain the normal kidney physiology^54^. Consequently, Talin-1 cleavage decouples integrins to actin cytoskeleton resulting in podocyte injury and kidney pathology, as previously described^39, 55–63^. Taking together, these data suggest that brain death specific proteolytic mechanisms may result in cleavage of cytoskeletal proteins, with SO donor kidneys affected more profoundly than GO.

Talin-1 is a specific Calpain target protein that can be cleaved on a number of sites by Calpain ^41, 64^, shown schematically in Figure 4C. As our results show that Talin-1 undergoes proteolytic processing in DBD kidneys and human kidney immortalized podocytes, we hypothesised that Talin-1 proteolysis resulted from activation of calcium-dependent protease Calpain and, as Calpain can cleave multiple targets, it could be a key proteolytic pathway in SO DBDs. This was supported by the PROTOMAP and western blot data on Calpain-1 specific proteolytic patterns indicative of enzymatic activation^42^.

Calpain proteases are activated directly by increased levels of intracellular levels of Ca^+2^, micromolar levels favors Calpain-1 activation^42^. Their physiological roles include cytoskeletal remodeling, cell cycle regulation and apoptosis in addition to proteolysis of specific targets^65^. Calpain-1 is known to proteolytically target Utrophin, α-actinin-4, and Dystroglycan^42^ and Calpain mediated degradation of Utrophin in muscle cells is reported to generate a ∼190kDa intermediate that has also been observed in the kidney PROTOMAP data^66^. To investigate a possible trigger for Calpain activation in DBD kidneys we built on our previous findings showing that TGF-β signaling is predominant in DBD kidneys compared to DCDs and enhanced in SO DBDs^23, 24^ As a result, we investigated if there was a direct link between increased TGF-β activity and Calpain activation in SO DBD kidneys.

We used *ex vivo* precision cut kidney slices and *in vitro* podocytes to show that TGF-β mediated Calpain-1 activation causing Talin proteolysis with degradation patterns that were fully aligned with the profile observed in DBD donor kidneys. More specifically, increasing TGF-β concentrations resulted in progressive Talin fragmentation and generation of the ∼125kDa proteoform that resembled the fragment detected in DBD kidneys. The role of Calpain in this cascade was confirmed by the protective effect of Calpeptin, a Calpain specific inhibitor. In our experiments, Calpeptin markedly reduced TGF-β driven degradation and prevented actin cytoskeletal dysregulation. This evidence of a TGF-β-Calpain interaction is supported by recent reports of TGF-β mediated Calpain activation during endothelial mesenchymal transition to myofibroblasts and onset of fibrosis; signaling events that were reversed by Calpeptin treatment^67, 68^. In the context of renal disease, Calpeptin has been demonstrated to prevent proteolytic processing of Talin and dysregulation of actin cytoskeleton, thus preventing glomerular injury, in rodent and *in vitro* models of experimental focal segmental glomerulosclerosis ^36, 69^. Our findings may provide a mechanistic insight to explain why transplants from living donors have superior function when compared to grafts procured from DBD donors.

Following severe cerebral injury, cerebral ischemia and brain stem death cause the release of endogenous cytokines and catecholamines that trigger a “sympathetic or catecholamine storm” ^13, 70, 71^. Increased levels of catecholamines in circulation increase the heart load and oxygen consumption with the production of reactive species at the cellular level. Hypoxic periods of kidney hypoperfusion associate with low energy delivery and depletion of ATP synthesis causing mitochondria disruption^9, 12^. These biological stressors trigger various cell stimuli such as membrane depolarization causing increased intracellular Ca^2+^ influx ^72^. In podocytes, impaired Ca^2+^ homeostasis, detected by specialized Ca^2+^-sensing proteins deciphers signalling cascades activate the Calpain network of endopeptidases. Once Calpain is activated it triggers proteolytic pathways that may modify the integrity of the podocyte cytoskeletal matrix in donor kidneys. These cytoskeletal changes may be sustained post-transplant, thereby contributing to progressive development of allograft dysfunction^73^.

Further studies would be informative to assess the impact of brain death triggered proteolysis on donor organs other than the kidney. It is highly probable that the complex biological changes during brain death and donor management may activate additional proteolytic mechanisms that would be important to further investigate. Moreover, the overall severity of cytoskeletal proteolytic degradation may also define whether the donor related injury is irreversible, possibly explaining the more limited protein cleavage that was observed in good outcome donor kidneys.

Emerging interest in the therapeutic application of Calpain inhibition to treat diseases such as muscular dystrophy, neurological injury, ischemia/reperfusion injury, clearly points towards preventative opportunities to improve donor grafts early after brain death^74, 75^. A targeted intervention to modulate or inhibit Calpain activation as a potential therapeutic strategy to prevent podocyte damage might not only improve allograft function but also survival, closing the gap between brain death and living donor outcomes.

As our observations were derived from donor kidneys, we were faced with limitations related to biological heterogeneity, reflected by the observation that in some donors Talin detection was low. This might also be explained by heterogeneity in the number of glomeruli in the analyzed preimplantation biopsies. Notably, our findings make associations of the podocyte alterations to allograft dysfunction although we need to acknowledge that kidney injury can extend to substructures beyond glomeruli.

To enhance these promising results, preclinical studies will be needed to confirm the direct interaction of TGF-β to Calpain activation and explore parallel proteolytic pathways that may also play a role in destabilizing the podocyte cytoskeleton.

In summary, our data, obtained from a unique cohort of biopsies from donor kidneys that developed long-term allograft dysfunction, suggests that DBD kidneys, when compared to DCD and LD, are more susceptible to protein degradation and that enhanced proteolysis can be linked to impaired graft function 12-month post-transplant. Employing complementary models of precision cut human kidney slices and human immortalized podocyte cells we provided initial evidence of the role of TGF-β and Calpain-1 in driving degradation and, importantly, we demonstrated *in-vitro* that podocyte Talin-1 degradation mirrored Talin proteolytic patterns identified in DBD kidney biopsies. Our findings suggest a scope to further study this alternative pathway of donor kidney injury that may offers novel therapeutic strategies to prevent podocyte damage preserving kidney function.

## Data Availability

http://proteomecentral.proteomexchange.org/cgi/GetDataset?ID=PXD022074

## AUTHOR CONTRIBUTION

MEK conceptualized the study. RHV, GIW, RN, BMK, RJP, MEK designed the research. RHV, JCK, LKF, MLT performed the experimental work. GIV, RN, BMK, RJP, MEK analysed and interpreted the data. RHV, JHIL, BMK, RJP and MEK wrote the manuscript. All authors revised the final manuscript.

## ACKNOWLEDGEMENTS

We thank the UK QUOD Consortium and NHS Blood and Transplant UK Registry for the clinical samples and metadata analysed in this study. We thank Ms Sandrine Rendel, Dr Sergei Maslau and Mr Tomas Surik for their support on the QUOD sample selection, Dr Philip Charles, Dr Letizia Lo Faro for their expert input in the study.

We would like thank Mr. Michael Schou Jensen, Ms Gitte Skou and Ms Gitte Kall for their assistance on PCKSs experiments. We also would like to thank the surgeons at the Department of Urology, Aarhus University Hospital for providing human tissue samples. Mr Jonathan Dixey for his assistance in cell culture within NHSBT and members of the Discovery Proteomics Facility within the TDI Mass Spectrometry Laboratory for expert help with mass spectrometry analysis.

## SUPPLEMENTARY DATA

**Supplementary Data Fig. 1.**
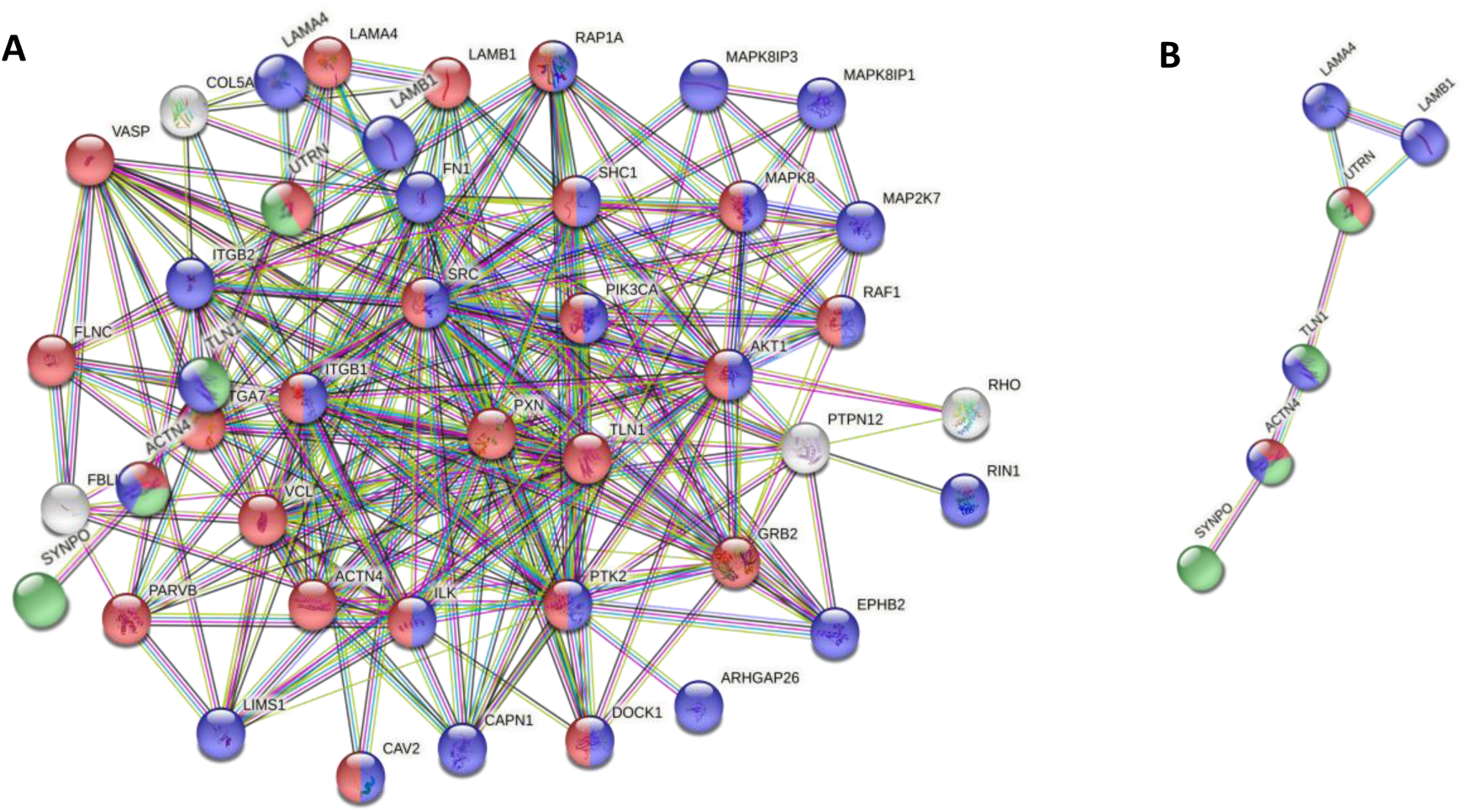
**A.** STRING pathway analysis of a subset of 65 cytoskeletal proteins show direct protein -protein interactions with enrichment of KEGG pathway focal adhesion (red) false discovery rate (FDR) 2.11 e-33, catabolic regulation (blue) FDR 1.06 e-09 and integrin mediated signaling (green) FDR 3.09e-9; **B.** Shortlisted proteins that show increased degradation in SO DBD kidneys formed a cluster of proteins mapped to kidney cortical cytoskeleton podocyte FDR 3.0 e-5.

**Supplementary Data Fig. 2.**
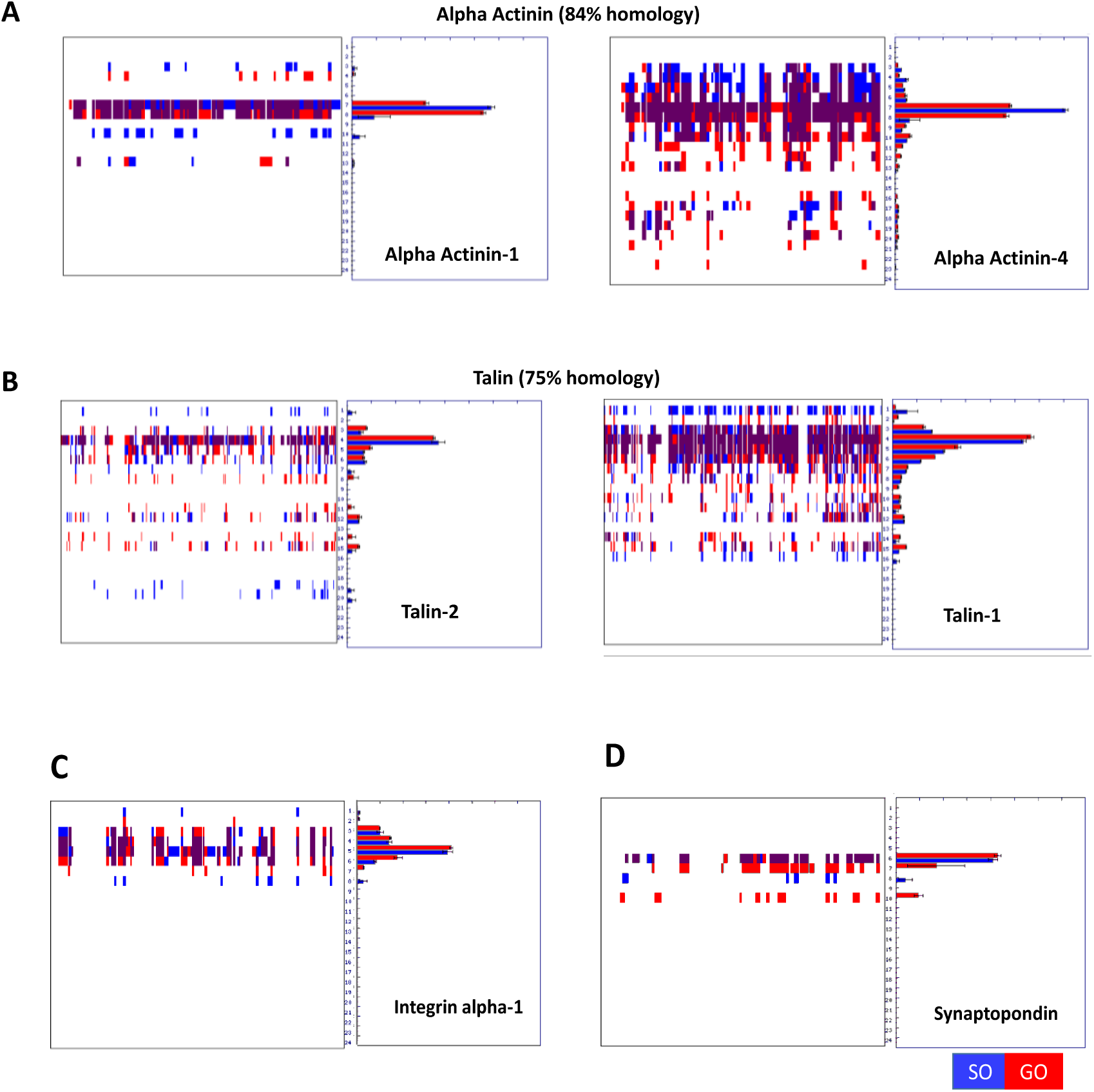
Degradation profiles. A&B Degradation profiles of protein isoforms detected in kidney biopsies. PROTOMAP peptographs of protein isoforms detected in brain death donor kidneys with suboptimal transplantation outcomes (red bars) compared to biopsies obtained from DBD kidneys with suboptimal transplantation outcomes (blue bars). Degradation profiles are shown from N- terminus (left) to C- terminus (right). **A.** Peptographs of α-actinin depict protein isoforms α-actinin-1 and α-actinin-4 that share 84% homology. Western blot analysis by α-actinin-4 confirmed protein proteolysis. **B.** Peptographs of Talin depict protein isoforms Talin-1 and Talin-2 that share 74% homology. Western blot analysis confirmed that Talin-1 detected fragments appeared in both Talin peptographs. **C.** Degradational profile of Integrin alpha-1 **D.** Degradation profile of Synaptopodin

**Supplementary Data Fig. 3.**
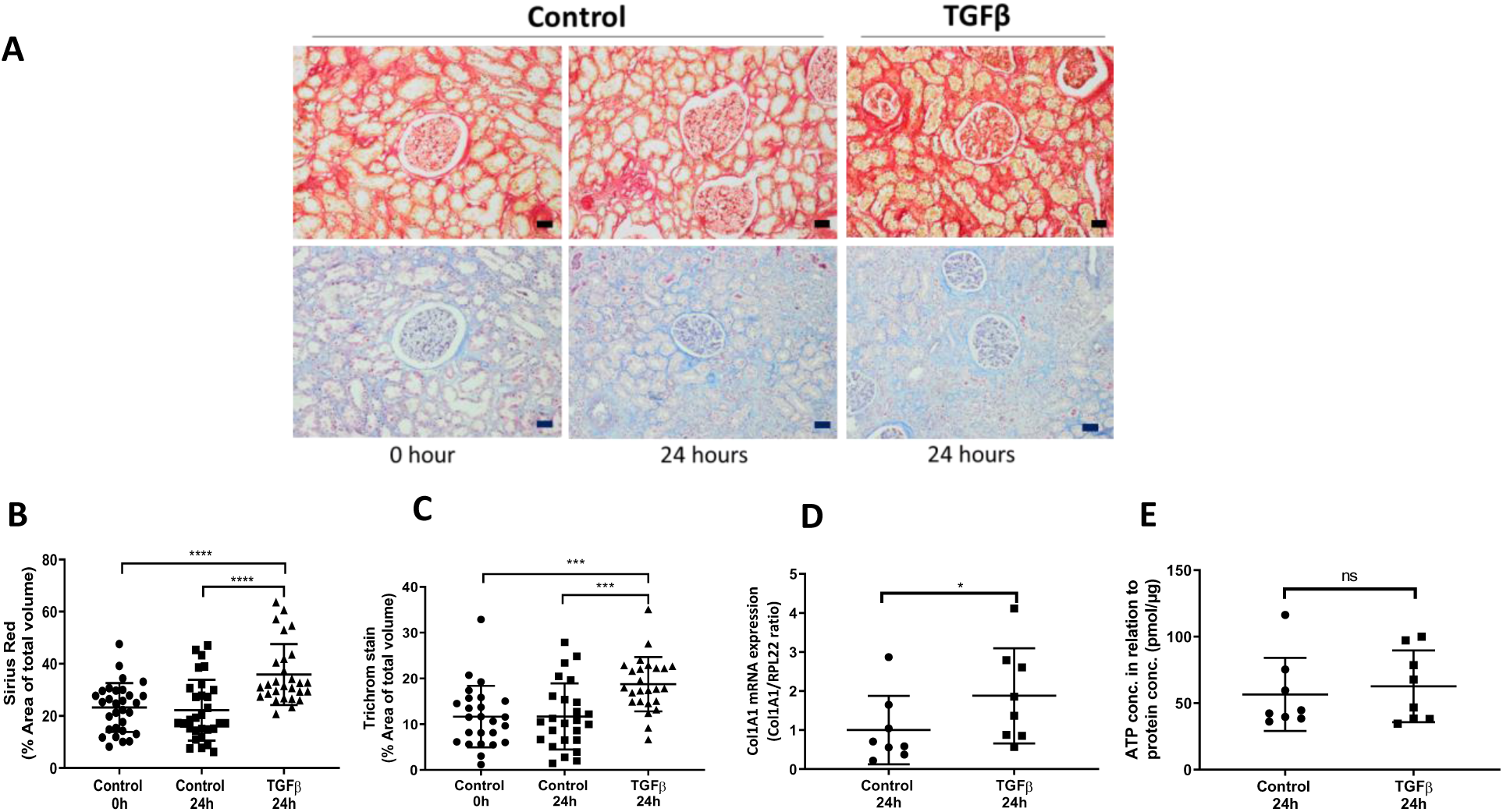
TG β t atm nt of h man p*recision-cut kidney slice (PCKS)* increases Calpain activity and causing pro-fibrotic morphological changes. **A.** PCKS stimulation with TGFβ (10ng) up to 24h caused the kidney slices to develop morphological changes; 20x magnification, 20µm scale bar. **B.** Sirius red and **C**. trichrome mason staining revealed that the PCKSs developed profibrotic histological characteristics of cortex, glomerular and tubular structures *(*P<0.05; t-test)* following TGFβ up to 24h **D.** Collagen1A1 mRNA expression is increased after TGFβ treatment of PCKS for 24h *(*P<0.05; t-test)*. **E.** Viability of the slices after treatment assessed by ATP content of the slices and confirmed that was affected as ATP measurement.

## REFERENCES

1. Oniscu GC, Ravanan R, Wu D, et al. Access to Transplantation and Transplant Outcome Measures (ATTOM): study protocol of a UK wide, in-depth, prospective cohort analysis. BMJ Open. 2016;6(2):e010377. doi:10.1136/bmjopen-2015-010377

2. Oniscu GC, Brown H, Forsythe JLR. Impact of cadaveric renal transplantation on survival in patients listed for transplantation. J Am Soc Nephrol. 2005;16(6):1859–1865. doi:10.1681/ASN.2004121092

3. Axelrod DA, Schnitzler MA, Xiao H, et al. An economic assessment of contemporary kidney transplant practice. Am J Transplant. 2018;18(5):1168–1176. doi:10.1111/ajt.14702

4. Iyamu Perisanidou L, Mumford L, Evans KM, et al. Causes of renal allograft failure in the UK: trends in UK Renal Registry and National Health Service Blood and Transplant data from 2000 to 2013. Nephrol Dial Transplant. 2018;34(2):355–364. doi:10.1093/ndt/gfy168

5. Westendorp WH, Leuvenink HG, Ploeg RJ. Brain death induced renal injury. Curr Opin Organ Transplant. 2011;16:151–156. doi:10.1097/MOT.0b013e328344a5dc

6. Bugge JF. Brain death and its implications for management of the potential organ donor. Acta Anaesthesiol Scand. 2009;53:1239–1250. doi:10.1111/j.1399-6576.2009.02064.x

7. UK Renal Registry. UK Renal Registry 21st Annual Report. 2019;3349. https://www.renalreg.org/publications-reports/.

8. Et DG, Martín C. Newsletter Transplant. 2018;23.

9. Van Erp AC, Rebolledo RA, Hoeksma D, et al. Organ-specific responses during brain death: increased aerobic metabolism in the liver and anaerobic metabolism with decreased perfusion in the kidneys. Sci Rep. 2018;8(1):4405. doi:10.1038/s41598-018-22689-9

10. Akhtar MZ, Sutherland AI, Huang H, Ploeg RJ, Pugh CW. The role of hypoxia-inducible factors in organ donation and transplantation: the current perspective and future opportunities. Am J Transplant. 2014;14(7):1481–1487. doi:10.1111/ajt.12737

11. Akhtar MZ, Huang H, Kaisar M, et al. Using an integrated -omics approach to identify key cellular processes that are disturbed in the kidney following brain death. Am J Transplant. November 2015. doi:10.1111/ajt.13626

12. Huang H, Van Dullemen LFA, Akhtar MZ, et al. Proteo-metabolomics reveals compensation between ischemic and non-injured contralateral kidneys after reperfusion. Sci Rep. 2018;8(1):1–12. doi:10.1038/s41598-018-26804-8

13. Schuurs T a., Morariu a. M, Ottens PJ, et al. Time-dependent changes in donor brain death related processes. Am J Transplant. 2006;6(August):2903–2911. doi:10.1111/j.1600-6143.2006.01547.x

14. Koudstaal LG, Ottens PJ, Ploeg RJ, Goor H Van, Leuvenink HGD. Brain Death Induces Inflammation in the Donor. 2008;(April):148–154. doi:10.1097/TP.0b013e31817ba53a

15. Bos EM, Leuvenink HGD, van Goor H, Ploeg RJ. Kidney grafts from brain dead donors: Inferior quality or opportunity for improvement? Kidney Int. 2007;72(7):797–805. doi:10.1038/sj.ki.5002400

16. Floerchinger B, Oberhuber R, Tullius SG. Effects of brain death on organ quality and transplant outcome. Transplant Rev. 2012;26(2):54–59. doi:10.1016/j.trre.2011.10.001

17. Meyer-Schwesinger C. The ubiquitin–proteasome system in kidney physiology and disease. Nat Rev Nephrol. 2019;15(7):393–411. doi:10.1038/s41581-019-0148-1

18. Meyer-Schwesinger C, Meyer TN, Münster S, et al. A new role for the neuronal ubiquitin C-terminal hydrolase-LI (UCH-LI) in podocyte process formation and podocyte injury in human glomerulopathies. J Pathol. 2009;217(September 2008):452–464. doi:10.1002/path.2446

19. Rinschen MM, Hoppe A-K, Grahammer F, et al. N-Degradomic Analysis Reveals a Proteolytic Network Processing the Podocyte Cytoskeleton. J Am Soc Nephrol. 2017;28(10):2867–2878. doi:10.1681/ASN.2016101119

20. Rinschen MM, Huesgen PF, Koch RE. The podocyte protease web: Uncovering the gatekeepers of glomerular disease. Am J Physiol - Ren Physiol. 2018;315(6):F1812–F1816. doi:10.1152/ajprenal.00380.2018

21. Fukasawa H. The role of the ubiquitin-proteasome system in kidney diseases. Clin Exp Nephrol. 2012;16:507–517. doi:10.1007/s10157-012-0643-1

22. Dix MM, Simon GM, Cravatt BF. Global Mapping of the Topography and Magnitude of Proteolytic Events in Apoptosis. Cell. 2008;134(4):679–691. doi:10.1016/j.cell.2008.06.038

23. Kaisar M, van Dullemen L, Charles P, et al. Subclinical Changes in Deceased Donor Kidney Proteomes are Associated with 12-month Allograft Function Posttransplantation – a Preliminary Study. Transplantation. 2018:1. doi:10.1097/TP.0000000000002358

24. De Kok MJ, McGuinness D, Shiels PG, et al. The neglectable impact of delayed graft function on long-term graft survival in kidneys donated after circulatory death associates with superior organ resilience. Ann Surg. 2019;XX(Xx):1-8. doi:10.1097/SLA.0000000000003515

25. https://www.nds.ox.ac.uk/research/quod.

26. Dix MM, Simon GM, Wang C, Okerberg E, Patricelli MP, Cravatt BF. Functional Interplay between Caspase Cleavage and Phosphorylation Sculpts the Apoptotic Proteome. Cell. 2012;150(2):426–440. doi:10.1016/j.cell.2012.05.040

27. Kaisar M, van Dullemen LFA, Thézénas M-L, et al. Plasma degradome affected by variable storage of human blood. Clin Proteomics. 2016;13:26. doi:10.1186/s12014-016-9126-9

28. Fischer R, Trudgian DC, Wright C, et al. Discovery of candidate serum proteomic and metabolomic biomarkers in ankylosing spondylitis. Mol Cell Proteomics. 2012;11(2):M111.013904. doi:10.1074/mcp.M111.013904

29. Simon GM, Dix MM, Cravatt BF. Comparative assessment of large-scale proteomic studies of apoptotic proteolysis. ACS Chem Biol. 2009;4(6):401–408. doi:10.1021/cb900082q

30. Niessen S, Hoover H, Gale AJ. Proteomic analysis of the coagulation reaction in plasma and whole blood using PROTOMAP. Proteomics. 2011;11(12):2377–2388. doi:10.1002/pmic.201000674

31. Kaisar M, van Dullemen LFA, Thézénas ML, Charles PD, Pleog RJ KB. Plasma Biomarker Profile Altered Through Variable Blood Storage. Clin Chem. 2016;62(9):1272–1277.

32. Mi H, Muruganujan A, Huang X, et al. Protocol Update for large-scale genome and gene function analysis with the PANTHER classification system (v.14.0). Nat Protoc. 2019;14(3):703–721. doi:10.1038/s41596-019-0128-8

33. Szklarczyk D, Gable AL, Lyon D, et al. STRING v11: Protein-protein association networks with increased coverage, supporting functional discovery in genome-wide experimental datasets. Nucleic Acids Res. 2019;47(D1):D607–D613. doi:10.1093/nar/gky1131

34. Jensen MS, Mutsaers HAM, Tingskov SJ, et al. Activation of the prostaglandin E 2 EP 2 receptor attenuates renal fibrosis in unilateral ureteral obstructed mice and human kidney slices. Acta Physiol. 2019;(February):e13291. doi:10.1111/apha.13291

35. Saleem MA, Hare MJO, Reiser J, et al. A Conditionally Immortalized Human Podocyte Cell Line Demonstrating Nephrin and Podocin Expression. JASN. 2002;(8):630–638.

36. Farmer LK, Rollason R, Whitcomb DJ, et al. TRPC6 Binds to and Activates Calpain, Independent of Its Channel Activity, and Regulates Podocyte Cytoskeleton, Cell Adhesion, and Motility. J Am Soc Nephrol. 2019:ASN.2018070729. doi:10.1681/ASN.2018070729

37. Dix MM, Simon GM, Cravatt BF. Global identification of caspase substrates using PROTOMAP (protein topography and migration analysis platform). Methods Mol Biol. 2014;1133:61–70. doi:10.1007/978-1-4939-0357-3_3

38. Feng D, DuMontier C, Pollak MR. The role of alpha-actinin-4 in human kidney disease. Cell Biosci. 2015;5:44. doi:10.1186/s13578-015-0036-8

39. Tian X, Kim JJ, Monkley SM, et al. Podocyte-associated talin1 is critical for glomerular filtration barrier maintenance. J Clin Invest. 2014;124(3):1098–1113. doi:10.1172/JCI69778

40. Moser M, Legate KR, Zent R, Fässler R. The tail of integrins, talin, and kindlins. Science. 2009;324(May):895–899. doi:10.1126/science.1163865

41. Franco SJ, Rodgers MA, Perrin BJ, et al. Calpain-mediated proteolysis of talin regulates adhesion dynamics. Nat Cell Biol. 2004;6(10):977–983. doi:10.1038/ncb1175

42. Goll DE, Thompson VF, Li H, Wei W, Cong J. The calpain system. Physiol Rev. 2003;83(3):731–801. doi:10.1152/physrev.00029.2002

43. Gabrijelcic-Geiger D, Mentele R, Meisel B, et al. Human μ-calpain: Simple isolation from erythrocytes and characterization of autolysis fragments. Biol Chem. 2001;382(12):1733–1737. doi:10.1515/BC.2001.209

44. Abdul-Hussien H, Soekhoe RGV, Weber E, et al. Collagen degradation in the abdominal aneurysm: A conspiracy of matrix metalloproteinase and cysteine collagenases. Am J Pathol. 2007;170(3):809–817. doi:10.2353/ajpath.2007.060522

45. Kaminska D, Tyran B, Mazanowska O, et al. Cytokine gene expression in kidney allograft biopsies after donor brain death and ischemia-reperfusion injury using in situ reverse-transcription polymerase chain reaction analysis. Transplantation. 2007;84(9):1118–1124. doi:10.1097/01.tp.0000287190.86654.74

46. Edward Sharplesa, Anna Casulab CB. UK Renal Registry 19th Annual Report: Chapter 3 Demographic and Biochemistry Profile of Kidney Transplant Recipients in the UK in 2015: National and Centre-specific Analyses. Nephron. 2016;132(1):99–110. doi:10.1159/000444818

47. Vaisar T, Hu JH, Airhart N, et al. Parallel Murine and Human Plaque Proteomics Reveals Pathways of Plaque Rupture. Circ Res. 2020:997–1022. doi:10.1161/circresaha.120.317295

48. Silva LM, Kryza T, Stoll T, et al. Integration of Two In-depth Quantitative Proteomics Approaches Determines the Kallikrein-related Peptidase 7 (KLK7) Degradome in Ovarian Cancer Cell Secretome. Mol Cell Proteomics. 2019;18(5):818–836. doi:10.1074/mcp.RA118.001304

49. Coppo R. Proteasome inhibitors in progressive renal diseases. Nephrol Dial Transplant. 2014;29(SUPPL. 1):25–30. doi:10.1093/ndt/gft271

50. Nagata M. Podocyte injury and its consequences. Kidney Int. 2016;89(6):1221–1230. doi:10.1016/j.kint.2016.01.012

51. Beeken M, Lindenmeyer MT, Blattner SM, et al. Alterations in the ubiquitin proteasome system in persistent but not reversible proteinuric diseases. J Am Soc Nephrol. 2014;25(11):2511–2525. doi:10.1681/ASN.2013050522

52. Hayek SS, Sever S, Ko Y-A, et al. Soluble Urokinase Receptor and Chronic Kidney Disease. N Engl J Med. 2015;373(20):1916–1925. doi:10.1056/NEJMoa1506362

53. Sever S, Altintas MM, Nankoe SR, et al. Proteolytic processing of dynamin by cytoplasmic cathepsin L is a mechanism for proteinuric kidney disease. J Clin Invest. 2007;117(8):2095–2104. doi:10.1172/JCI32022

54. Calderwood D a, Campbell ID, Critchley DR. Talins and kindlins: partners in integrin-mediated adhesion. Nat Rev Mol Cell Biol. 2013;14(8):503–517. doi:10.1038/nrm3624

55. Lennon R, Randles MJ, Humphries MJ. The importance of podocyte adhesion for a healthy glomerulus. Front Endocrinol (Lausanne). 2014;5(OCT):1–17. doi:10.3389/fendo.2014.00160

56. Lennon R, Byron A, Humphries JD, et al. Global Analysis Reveals the Complexity of the Human Glomerular Extracellular Matrix. J Am Soc Nephrol. 2014;25(5):939–951. doi:10.1681/ASN.2013030233

57. Faul C, Asanuma K, Yanagida-Asanuma E, Kim K, Mundel P. Actin up: regulation of podocyte structure and function by components of the actin cytoskeleton. Trends Cell Biol. 2007;17(9):428–437. doi:10.1016/j.tcb.2007.06.006

58. Henderson JM, Alexander MP, Pollak MR. Patients with ACTN4 mutations demonstrate distinctive features of glomerular injury. J Am Soc Nephrol. 2009;20(5):961–968. doi:10.1681/ASN.2008060613

59. Mundel P, Shankland SJ. Podocyte biology and response to injury. J Am Soc Nephrol. 2002;13(12):3005–3015. http://www.ncbi.nlm.nih.gov/pubmed/12444221. Accessed August 3, 2016.

60. Gu C, Yaddanapudi S, Weins A, et al. Direct dynamin-actin interactions regulate the actin cytoskeleton. EMBO J. 2010;29(21):3593–3606. doi:10.1038/emboj.2010.249

61. Raats CJ, van den Born J, Bakker MA, et al. Expression of agrin, dystroglycan, and utrophin in normal renal tissue and in experimental glomerulopathies. Am J Pathol. 2000;156(5):1749–1765. doi:10.1016/S0002-9440(10)65046-8

62. Tadokoro S, Shattil SJ, Eto K, et al. Talin binding to integrin β tails: A final common step in integrin activation. Science (80-). 2003;302(5642):103–106. doi:10.1126/science.1086652

63. Calderwood DA, Ginsberg MH. Talin forges the links between integrins and actin. Nat Cell Biol. 2003;5(8):694–697. doi:10.1038/ncb0803-694

64. Rees DJ, Ades SE, Singer SJ, Hynes RO. Sequence and domain structure of talin. Nature. 1990;347(6294):685–689. doi:10.1038/347685a0

65. Moldoveanu T, Hosfield CM, Lim D, Jia Z, Davies PL. Calpain silencing by a reversible intrinsic mechanism. Nat Struct Biol. 2003;10(5):371–378. doi:10.1038/nsb917

66. Courdier-Fruh I, Briguet A. Utrophin is a calpain substrate in muscle cells. Muscle Nerve. 2006;33(6):753–759. doi:10.1002/mus.20549

67. Kim DH, Beckett JD, Nagpal V, et al. Calpain 9 as a therapeutic target in TGFβ-induced mesenchymal transition and fibrosis. Sci Transl Med. 2019;11(501):eaau2814. doi:10.1126/scitranslmed.aau2814

68. W.-J. T, Q.-Y. T, T. W, M. L, L. Z, Z.-S. C. Calpain 1 regulates TGF-β1-induced epithelial-mesenchymal transition in human lung epithelial cells via PI3K/Akt signaling pathway. Am J Transl Res. 2017;9(3):1402–1409. http://www.embase.com/search/results?subaction=viewrecord&from=export&id=L615094474.

69. Verheijden KAT, Sonneveld R, Bakker-van Bebber M, Wetzels JFM, van der Vlag J, Nijenhuis T. The Calcium-Dependent Protease Calpain-1 Links TRPC6 Activity to Podocyte Injury. J Am Soc Nephrol. 2018;(464):ASN.2016111248. doi:10.1681/ASN.2016111248

70. Murugan R, Venkataraman R, Wahed AS, et al. Increased plasma interleukin-6 in donors is associated with lower recipient hospital-free survival after cadaveric organ transplantation. Crit Care Med. 2008;36(6):1810–1816. doi:10.1097/CCM.0b013e318174d89f

71. Skrabal CA, Thompson LO, Potapov E V., et al. Organ-specific regulation of pro-inflammatory molecules in heart, lung, and kidney following brain death. J Surg Res. 2005;123(1):118–125. doi:10.1016/j.jss.2004.07.245

72. Rafaela Bagur1 and György Hajnóczky. Intracellular Ca 2 + Sensing : Its Role in Calcium Homeostasis and Signaling. Cell. 2017. doi:10.1016/j.molcel.2017.05.028

73. Jaiswal. Regulation of podocyte actin dynamics by calcium. Bone. 2014;23(1):1–7. doi:10.1016/j.semnephrol.2012.06.003.Regulation

74. Ono Y, Saido TC, Sorimachi H. Calpain research for drug discovery: Challenges and potential. Nat Rev Drug Discov. 2016;15(12):854–876. doi:10.1038/nrd.2016.212

75. Neuhof C. Calpain system and its involvement in myocardial ischemia and reperfusion injury. World J Cardiol. 2014;6(7):638. doi:10.4330/wjc.v6.i7.638

